# Efficacy and safety of emapalumab in macrophage activation syndrome

**DOI:** 10.1101/2022.12.12.22283141

**Authors:** Fabrizio De Benedetti, Alexei A Grom, Paul Brogan, Claudia Bracaglia, Manuela Pardeo, Giulia Marucci, Despina Eleftheriou, Charalampia Papadopoulou, Grant S Schulert, Pierre Quartier, Jordi Antón, Christian Laveille, Rikke Frederiksen, Veronica Asnaghi, Maria Ballabio, Philippe Jacqmin, Cristina de Min

## Abstract

**Objectives:** Macrophage activation syndrome (MAS) is a severe, life-threatening complication of systemic juvenile idiopathic arthritis (sJIA) and adult onset Still’s disease (AOSD). The objective of this study was to confirm the adequacy of an emapalumab dosing regimen in relation to interferon-γ (IFNγ) activity by assessing efficacy and safety. The efficacy outcome was MAS remission by week 8, based on clinical and laboratory criteria.

**Methods:** We studied emapalumab, a human anti–IFNγ antibody, administered with background glucocorticoids, in a prospective open-label, single-group trial involving patients who had MAS secondary to sJIA or AOSD and had previously failed high-dose glucocorticoids. The study foresaw 4-week treatment that could be shortened or prolonged based on investigator’s assessment of response. Patients could enter a long-term (12 months) follow-up study.

**Results:** Fourteen patients received emapalumab. All patients completed the trial, entered the long-term follow-up and were alive at the end of the follow-up. The investigated dosing regimen, based on an initial loading dose followed by maintenance doses, was appropriate, as shown by rapid neutralisation of IFNγ activity, demonstrated by prompt decrease in serum C-X-C motif chemokine ligand 9 (CXCL9) levels. By week 8, remission of MAS was achieved in 13 of the 14 patients at a median time of 25 days. Viral infections and positive viral tests were observed during the trial and during the long-term follow-up.

**Conclusions:** Neutralisation of IFNγ with emapalumab was efficacious in inducing remission of MAS secondary to sJIA or AOSD in patients who had failed high-dose glucocorticoids.

**KEY MESSAGES:** *What is already known on this topic?:* Macrophage activation syndrome (MAS) is a severe and potentially life-threatening complication of systemic juvenile idiopathic arthritis and adult-onset Still’s disease. There are no therapeutic options that have been prospectively investigated in MAS. Data in animal models and *ex vivo* data from humans with MAS led to the hypothesis that interferon-γ (IFNγ) has a pathogenic role in MAS.

*What does this study add?:* This open-label multicentre trial using emapalumab, an anti–IFNγ antibody, in patients who have failed to respond to high-dose glucocorticoids, demonstrates that IFNγ has a pathogenic role in MAS and that its neutralisation leads to MAS remission.

*How might this affect research, clinical practice or policy?:* The results of this study show that neutralisation of IFNγ with emapalumab is a therapeutic option for patients with severe MAS who have failed standard of care with high-dose glucocorticoids.

## INTRODUCTION

Macrophage activation syndrome (MAS) is a form of secondary haemophagocytic lymphohistiocytosis (HLH) occurring as a life-threatening complication of rheumatic diseases.[1] It is most frequent in systemic juvenile idiopathic arthritis (sJIA) and adult-onset Still’s disease (AOSD), affecting about 10–20% of patients.[1-4] sJIA and AOSD are considered to be the same disease named differently depending only on age at onset being below or above 16 years, respectively.[2,3,5] The incidence and features of MAS are similar in the two age groups.[2,5] We use the term sJIA/AOSD to collectively identify these patients.

Similar to other forms of HLH, MAS is caused by excessive activation and expansion of T lymphocytes and macrophages resulting in hyperinflammation.[1,6,7] MAS is characterised by fever, hepatosplenomegaly, cytopenias, liver dysfunction, coagulation abnormalities and hyperferritinaemia, and may progress to multiple organ failure, with mortality rates of 10–20%.[4]

MAS is treated with high-dose glucocorticoids with satisfactory response in two-thirds of the patients.[4,8] For patients unresponsive to glucocorticoids, ciclosporin is usually added. Other approaches, including cyclophosphamide, etoposide, intravenous immunoglobulin, etanercept, anakinra, tocilizumab, JAK inhibitors and plasmapheresis, have been described in case reports or small series.[4,8-12] None of these regimens have been prospectively investigated.

Overproduction of interferon-γ (IFNγ) is present and pathogenic in animal models of MAS.[13-16] In sJIA/AOSD, high IFNγ activity demonstrated by high serum levels of C-X-C motif chemokine ligand 9 (CXCL9), a chemokine exclusively induced by IFNγ,[17] is associated with MAS onset and severity.[18-22]

We studied emapalumab, a human anti-IFNγ antibody, in patients with MAS secondary to sJIA/AOSD who failed to respond to high-dose glucocorticoids.

## METHODS

This phase 2, open-label, single-arm trial (NI-0501-06; ClinicalTrials.gov Identifier NCT02069899) was conducted at five sites in Italy, France, Spain, the United Kingdom and the United States. It comprised screening, a treatment period of 28 days and a short-term follow-up of 4 weeks. Data were collected up to 12 months in a long-term follow-up study (NI-0501-05; ClinicalTrials.gov Identifier NCT03311854) (Fig. S1 in the Supplementary Appendix). Results from both studies are reported.

Approval was obtained from institutional review boards or independent ethics committees at each participating centre. All patients or their legal representatives provided written informed consent. FDB, AG, MB and CdM designed the study. The sponsor was also responsible for data collection, management and analysis. All authors vouch for the accuracy and completeness of the data and analyses, and for the fidelity of the study to the protocol.

### Patient involvement

The study was presented and discussed at the 2016 and 2017 meeting of the sJIA Foundation in Washington, DC, USA. Information on the study for patients and families was published on the sJIA Foundation website. Dissemination of the data gathered from the study is also planned to be discussed with the sJIA Foundation.

### Patients

Patients had sJIA based on International League Against Rheumatism criteria,[23] or AOSD based on Yamaguchi criteria.[24] For patients presenting with MAS at sJIA onset, presumption of sJIA was based on Childhood Arthritis and Rheumatology Research Alliance criteria.[25] Patients had MAS, according to American College of Rheumatology/European League Against Rheumatism criteria,[26] and an inadequate response to high-dose intravenous glucocorticoids according to the treating physicians. High-dose glucocorticoids were defined as ≥2 mg/kg/day of prednisone equivalent in two divided doses, or at least 60 mg/day in patients weighing 30 kg or more, including but not limited to pulses up to 30 mg/kg/day for at least 3 consecutive days. Patients with active infections potentially favoured by IFNγ neutralisation (typical and atypical mycobacteria, *Histoplasma capsulatum, Shigella, Salmonella, Campylobacter* and *Leishmania*) were excluded. Eligibility criteria are listed in Table S1.

### Treatment

Emapalumab infusions were administered on a background of glucocorticoids. The initial dose of emapalumab was 6 mg/kg on day 0, followed by 3 mg/kg every 3 days until day 15 and twice weekly until day 28. Treatment with emapalumab could be stopped upon investigator’s assessment of remission, but not before 3 doses of emapalumab have been administered. Frequency between infusions could be shortened, dose could be increased or treatment prolonged upon investigator’s assessment of unsatisfactory response. Ciclosporin could be continued, if started at least 3 days prior to initiating emapalumab. Inhibitors of IL-1 and IL-6 were not allowed. An amendment allowed continuation of anakinra, if started at least 3 days before initiation of emapalumab, and its introduction during the study, at a maximum dose of 4 mg/kg/day for the treatment of the underlying sJIA/AOSD. Tocilizumab and canakinumab were not allowed during the trial. Prophylaxis against herpes zoster was administered as per local standards. Glucocorticoid tapering could be initiated as soon as the patients’ conditions allowed based on investigator’s assessment.

### Outcomes

The objective was to confirm the adequacy of the emapalumab dosing regimen in relation to IFNγ activity by assessing efficacy and safety. Serum levels of emapalumab, total (free and emapalumab-bound) IFNγ, CXCL9 and soluble interleukin [IL]-2 receptor were measured (Methods in Supplementary Appendices 1 and 2).

The efficacy outcome was MAS remission by week 8, defined as resolution of clinical signs and symptoms (Table S2) according to the physician global assessment (visual analogue scale ≤1/10) and white blood cell and platelet count above lower limit of normal, lactate dehydrogenase (LDH), alanine aminotransferase and aspartate aminotransferase below 1.5 times upper limit of normal (ULN), fibrinogen greater than 100 mg/dL and ferritin levels [27] decreased by at least 80% or below 2000 ng/mL, whichever was lower. Other efficacy evaluations included glucocorticoid dose (expressed as mg/kg/day of prednisone-equivalent) and survival. Adverse events (AEs) were assessed. During the long-term follow-up, evaluations included MAS episodes, AEs, pharmacokinetics and pharmacodynamics.

### Statistical analysis

The analysis population included all patients. Categorical variables are presented with the number and percentage within each category. Continuous variables are reported as median (range). Laboratory parameters of MAS were measured locally.

## RESULTS

### Study population

Fourteen patients received emapalumab. All completed the study and entered long-term follow-up. Thirteen patients had sJIA onset before 16 years, and one AOSD with onset at 16 years and 9 months. Four patients had presumption of sJIA, which was later confirmed. Six patients had a total of 19 previous MAS episodes in their history, treated with high-dose glucocorticoids, with the addition of anakinra and/or ciclosporin in approximately half.

During the week preceding emapalumab, all patients were receiving high-dose intravenous glucocorticoids, as per protocol definition. In addition, eight were receiving ciclosporin and seven anakinra (Table 1). Of the patients treated with anakinra, three were receiving standard doses for sJIA (≤4.0 mg/kg/day), and four high doses ranging from 7.5 to 15 mg/kg/day. Patients had severe MAS at baseline, shown by high physician assessment of MAS activity and marked abnormalities of laboratory parameters (Table 2). Notably, worsening or no improvement of laboratory parameters was observed between screening and baseline, despite the ongoing treatment.

**Table 1.**
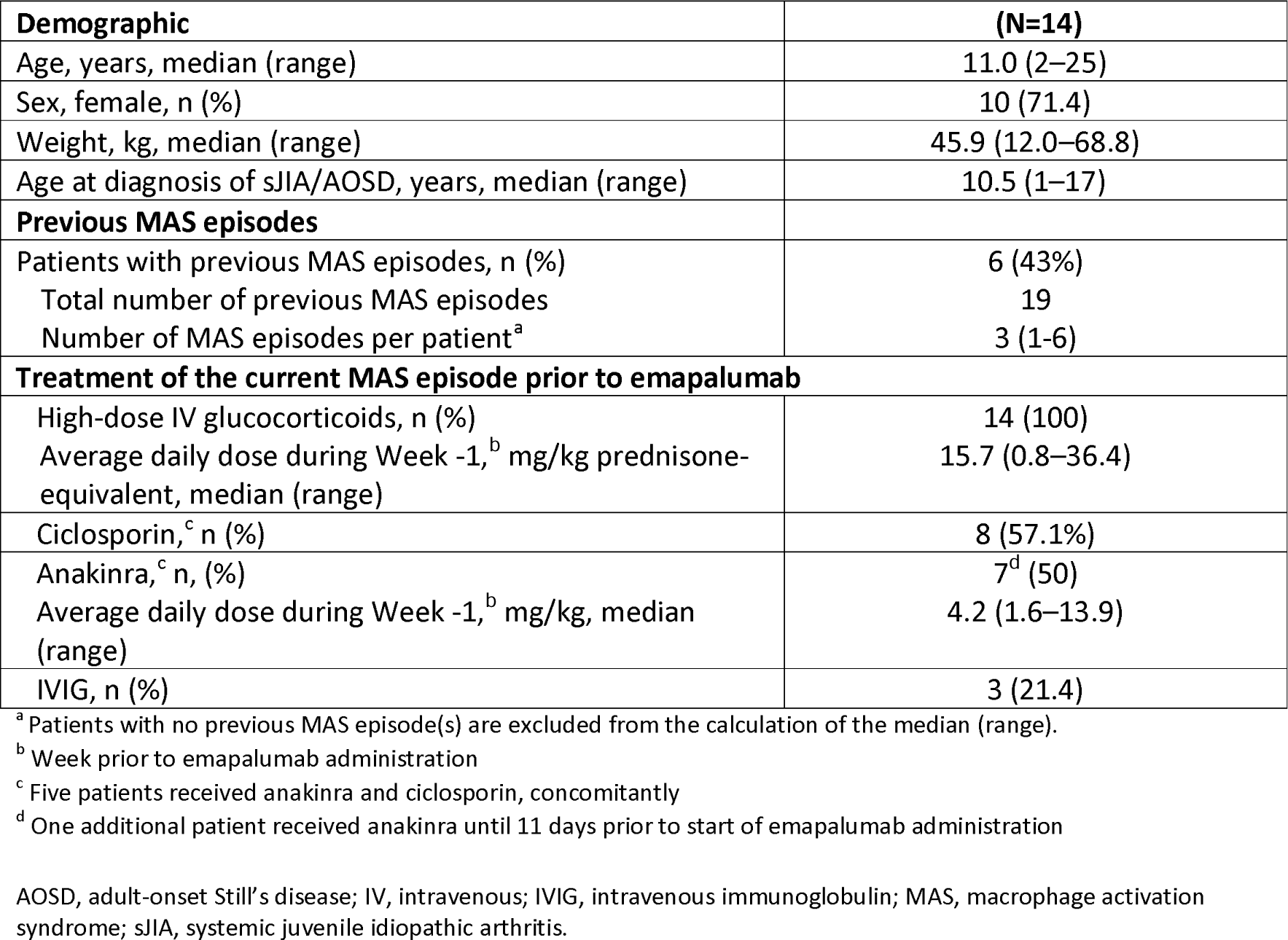
Baseline demographics and clinical characteristics of the patients.

| **Demographic** | **(N=14)** |
| --- | --- |
| Age, years, median (range) | 11.0 (2–25) |
| Sex, female, n (%) | 10 (71.4) |
| Weight, kg, median (range) | 45.9 (12.0–68.8) |
| Age at diagnosis of sJIA/AOSD, years, median (range) | 10.5 (1–17) |
| **Previous MAS episodes** |  |
| Patients with previous MAS episodes, n (%) | 6 (43%) |
| Total number of previous MAS episodes | 19 |
| Number of MAS episodes per patient<sup>a</sup> | 3 (1–6) |
| **Treatment of the current MAS episode prior to emapalumab** |  |
| High-dose IV glucocorticoids, n (%) | 14 (100) |
| Average daily dose during Week -1, <sup>b</sup> mg/kg prednisone-equivalent, median (range) | 15.7 (0.8–36.4) |
| Ciclosporin, <sup>c</sup> n (%) | 8 (57.1%) |
| Anakinra, <sup>c</sup> n, (%) | 7<sup>d</sup> (50) |
| Average daily dose during Week -1, <sup>b</sup> mg/kg, median (range) | 4.2 (1.6–13.9) |
| IVIg, n (%) | 3 (21.4) |

<sup>a</sup> Patients with no previous MAS episode(s) are excluded from the calculation of the median (range).
<sup>b</sup> Week prior to emapalumab administration
<sup>c</sup> Five patients received anakinra and ciclosporin, concomitantly
<sup>d</sup> One additional patient received anakinra until 11 days prior to start of emapalumab administration
AOSD, adult-onset Still's disease; IV, intravenous; IVIg, intravenous immunoglobulin; MAS, macrophage activation syndrome; sJIA, systemic juvenile idiopathic arthritis.

**Table 2.** MAS activity, as defined by physician’s assessment, and laboratory features of MAS before, during and after emapalumab treatment.

|  | Screening (N=14) | Baseline (N=14) | Week 2 (N=14) | Week 4 (N=14) | Week 8 (N=14) |
| --- | --- | --- | --- | --- | --- |
| Physician assessment of MAS activity<sup>a</sup>,<br>centimeter, median (range) | 8.3 (2.0–10.0) | 8.0 (2.0–10.0) | 2.3 (0.0–9.5) | 0.3 (0.0–6.5) | 0 (0.0–1.0) |
| Ferritin, ng/mL, median (range) | 19,865 (367–192,584) | 25,709 (716–192,584) | 1,410 (65–29,099) | 180 (22–18,430) | 55 (4–561) |
| Increased, n (%)<sup>b</sup> | 13 (93) | 14 (100) | 9 (64) | 2 (14) | 0 (0) |
| WBCs, 10<sup>9</sup>/L, median (range) | 5.8 (1.0–25.7) | 5.1 (0.9–25.7) | 8.4 (0.9–32.6) | 12.3 (2.4–34.7) | 9.5 (4.8–22.6) |
| Decreased, n (%)<sup>c</sup> | 6 (43) | 7 (50) | 4 (29) | 1 (7) | 1 (7) |
| Platelets, 10<sup>9</sup>/L, median (range) | 152 (48–546) | 112 (52–558) | 284 (55–590) | 336 (104–591) | 383 (214–915) |
| Decreased, n (%)<sup>c</sup> | 6 (43) | 9 (64) | 4 (29) | 2 (14) | 0 (0) |
| Aspartate aminotransferase, U/L, median (range) | 244 (26–3814) | 190 (19–2039) | 34 (5–146) | 29 (16–100) | 28 (15–52) |
| Increased, n (%)<sup>d</sup> | 12 (86) | 10 (71) | 3 (21) | 1 (7) | 0 (0) |
| Alanine aminotransferase, U/L, median (range) | 190 (139–3179) | 302 (70–1492) | 61 (18–327) | 45 (23–195) | 28 (7–59) |
| Increased, n (%)<sup>d</sup> | 14 (100) | 14 (100) | 6 (43) | 5 (35.7) | 0 (0) |
| LDH, U/L, median (range) | 1,699 (859–12,734) | 1,502 (588–12,734) | 518 (283–1572) | 592 (277–810) | 497 (306–741) |
| Increased, n (%)<sup>d</sup> | 13 (100) | 13 (93) | 6 (43) | 4 (29) | 2 (14) |
| Fibrinogen, mg/dL, median (range) | 230 (137–341) | 174 (70–465) | 244 (145–515) | 279 (200–460) | 337 (190–460) |
| Decreased, n (%)<sup>e</sup> | 0 (0) | 1 (7) | 0 (0) | 0 (0) | 0 (0) |

<sup>a</sup> MAS activity was based on a 10-cm visual analogue scale, with scores ranging from 0 to 10 and higher score indicating higher activity.
<sup>b</sup> Increased ferritin was defined as ferritin >684 µg/L as per 2016 Classification Criteria for MAS complicating sJIA. [23]
<sup>c</sup> Decreased white blood cell and platelet were defined as counts below the lower limit of normal for age of local laboratory
<sup>d</sup> Increased alanine aminotransferase, aspartate aminotransferase and LDH were defined as values above 1.5 the upper limit of normal range of local laboratory.
<sup>e</sup> Decreased fibrinogen was defined as fibrinogen <100 mg/dL.
LDH, lactate dehydrogenase; MAS, macrophage activation syndrome; WBC, white blood cells.

Of the 14 patients, six received emapalumab up to day 28. In seven patients who had MAS remission per investigator’s assessment, it was possible to discontinue emapalumab earlier. One patient continued emapalumab up to day 39. The median duration of treatment was 27 days (range, 7–39) with the number of infusions ranging from 3 to 17 per patient.

### Pharmacokinetics and pharmacodynamics

All patients received an initial dose of emapalumab of 6 mg/kg followed by doses of 3 mg/kg. The initial dose allowed to rapidly reach serum concentrations of emapalumab close to the steady-state concentrations obtained with the doses of 3 mg/kg (Fig. 1A). The dosing interval was transiently shortened from 3 to 2 days in three patients based on clinical and laboratory parameters suggestive of incomplete response. In two of these patients the dosing interval was shortened for several infusions. In these two patients, measurement of emapalumab concentrations showed rapid drug clearance consistent with target-mediated drug disposition associated with high levels of total IFNγ (Fig. S2).[28]

**Figure 1.**
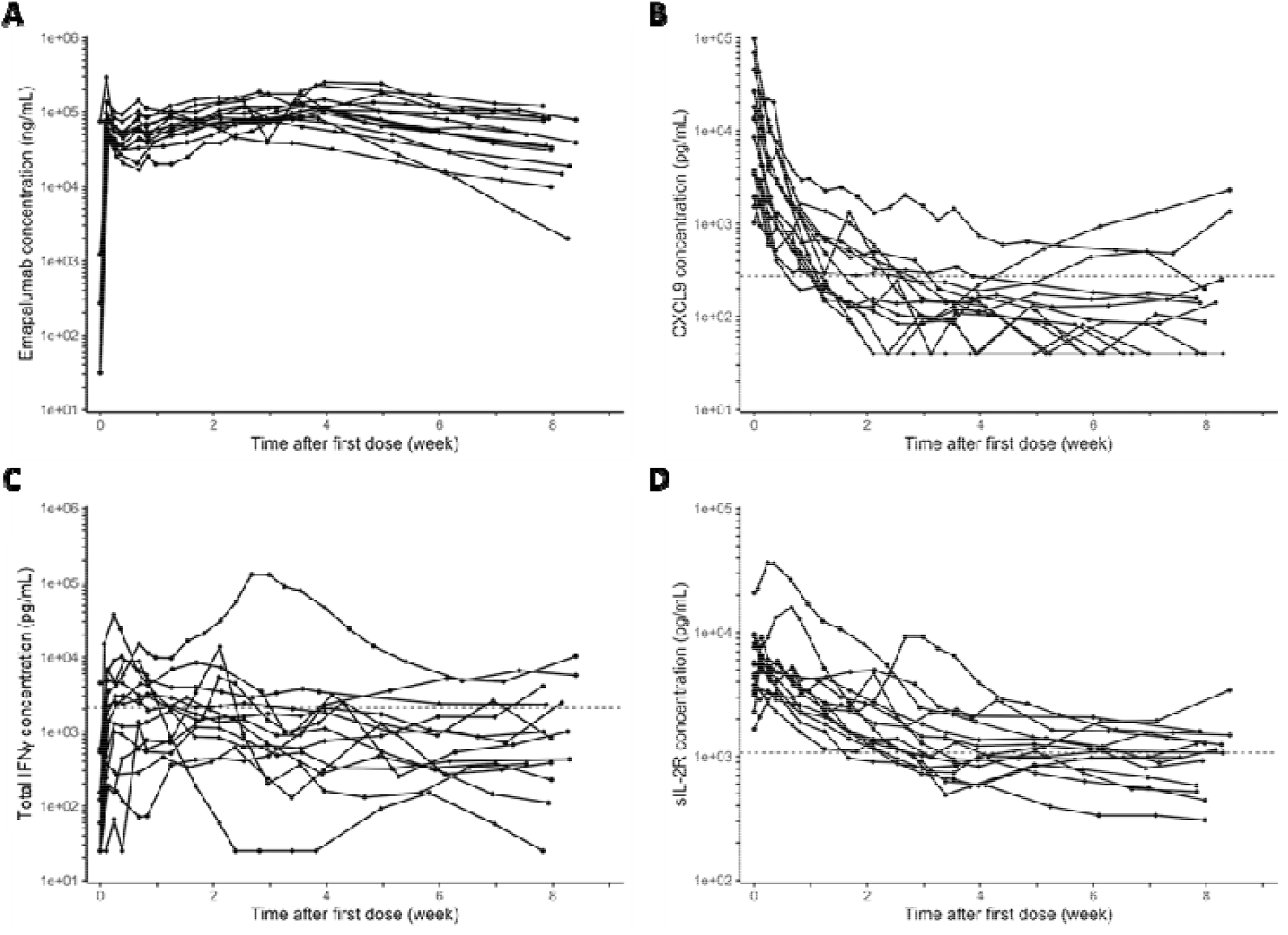
Pharmacokinetic (emapalumab^a^ [A]) and pharmacodynamic (CXCL9^b^ [B], total IFNγ ^c^ [C] and sIL-2R^d^ [D]) parameters in patients with MAS treated with emapalumab. ^a^ Serum emapalumab concentrations in patients treated with emapalumab (initial dose of 6 mg/kg, followed by intended maintenance dosing of 3 mg/kg every 3 days until day 15, and twice weekly until day 28). ^b^Serum concentrations of CXCL9, a biomarker of interferon-γ activity, being, at baseline, 4 to 362 times the 95^th^ percentile of healthy volunteers (271 pg/mL),[24] indicated by the horizontal dotted line. ^c^Total IFNγ (free and emapalumab-bound) that reflects IFNγ production, ranging, at day 3, from normal to 12 times the 95^th^ percentile of healthy volunteers receiving emapalumab (2084 pg/mL) indicated by the horizontal dotted line. ^d^Serum sIL-2R, a biomarker of T cell activation, being, at baseline, 1.6 to 20 times the 95^th^ percentile of healthy volunteers (1071 pg/mL, unpublished data) indicated by the horizontal dotted line. IFNγ, interferon-γ; MAS, macrophage activation syndrome; sIL-2R, soluble interleukin-2 receptor.

At baseline, CXCL9 concentrations were markedly elevated in all patients, indicating high IFNγ activity (Fig. 1B). Total IFNγ concentrations were variable across patients (Fig. 1C). Total IFNγ concentrations at day 3 (at equilibrium between free and emapalumab-bound IFNγ) reflect IFNγ production at baseline, as previously described.[27,28] There was no correlation between total IFNγ at day 3 and CXCL9 levels at baseline (not shown). Soluble IL-2 receptor levels at baseline were markedly elevated in all patients (Fig. 1D).

Emapalumab administration led to rapid decrease in serum CXCL9 levels, indicating neutralisation of IFNγ activity and therefore the appropriateness of the selected dosing regimen. Patients with elevated total IFNγ at day 3 also showed a decrease in total IFNγ concentrations over time indicating decreasing IFNγ production over time. Soluble IL-2 receptor concentrations also decreased markedly (Fig. 1D), indicating decreasing T cell activation. During long-term follow-up, in the absence of high IFNγ production, as shown by low levels of total IFNγ, emapalumab showed a slow linear terminal elimination phase with a half-life of 24 days, like that in healthy subjects (Fig. S3).[28] Total IFNγ, CXCL9 and soluble IL-2 receptor levels were close to, or within, the normal range (Table S3).

### Efficacy

During emapalumab administration, physician global assessment of MAS activity and MAS laboratory parameters rapidly improved in all patients while glucocorticoid were tapered.

By week 8, 13 patients (93%) achieved MAS remission at a median time of 25 days after emapalumab initiation, the earliest at day 9 (Fig. 2). At week 8, two of the 13 patients who had previously achieved MAS remission did not meet the criteria of MAS remission because of a single laboratory abnormality (one with LDH 1.7-fold above the ULN; 1 with white blood cells at 4.8×10^9^/L with lower limit of normal at 5.5×10^9^/L). One patient who stopped emapalumab after three doses upon investigator’s assessment of remission never met the criteria of MAS remission only because of LDH levels 1.5-fold above the ULN.

**Figure 2.**
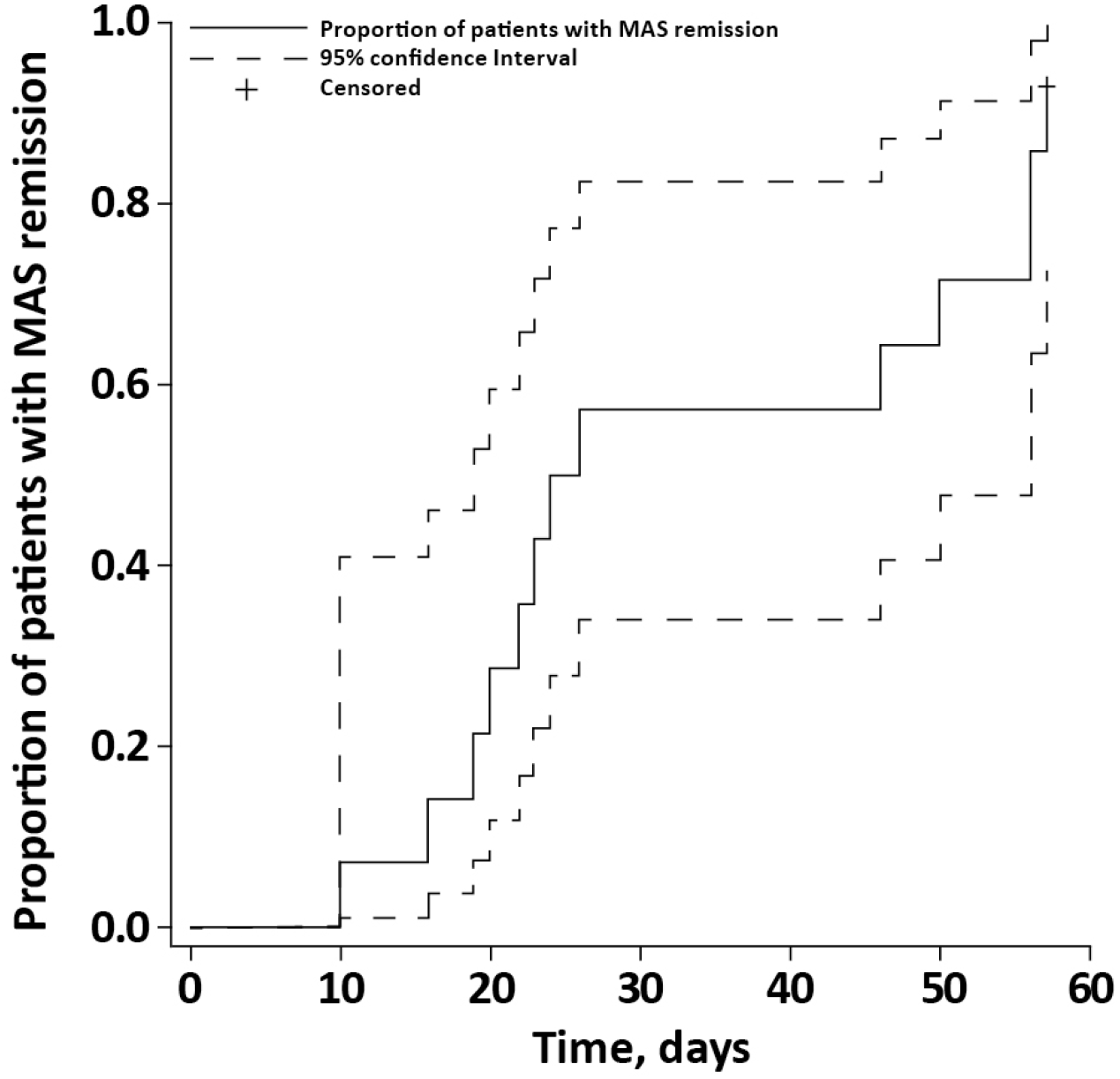
Time to MAS remission^a^. ^a^Time to MAS remission defined as resolution of clinical signs and symptoms according to the investigator (visual analog scale ≤1/10) (Table S2) and resolution of the abnormalities of MAS laboratory parameters: white blood cell and platelet count above lower limit of normal, LDH below 1.5 times ULN, alanine aminotransferase/aspartate aminotransferase below 1.5 times ULN, fibrinogen greater than 100 mg/dL and ferritin levels decreased by at least 80% or below 2000 ng/mL, whichever was lower. The continuous line represents the proportion of patients with MAS remission, the dotted line represents the 95% confidence interval. The cross indicates the censored patient. LDH, lactate dehydrogenase; MAS, macrophage activation syndrome; ULN, upper limit of normal.

All laboratory parameters of MAS rapidly improved upon initiation of emapalumab treatment with all parameters being normal in the majority of the patients by week 4 (Fig 3, Table 2 and Fig S4).

**Figure 3.**
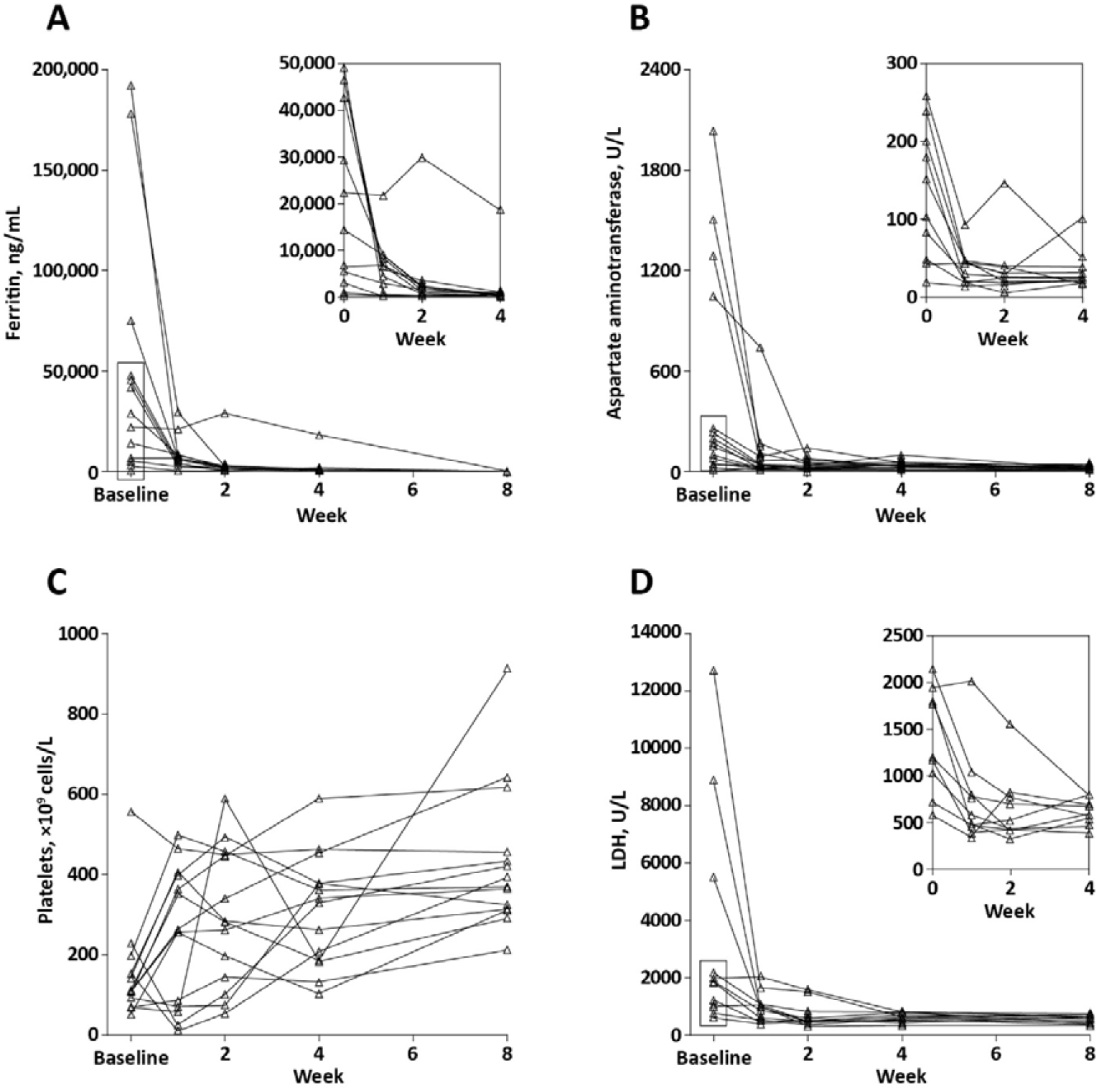
Changes in ferritin^a^ (A), aspartate aminotransferase^b^ (B), platelets (C) and LDH^c^ (D) levels over time. ^a^Panel A insert shows in detail changes from baseline to week 4 for patients with baseline levels of ferritin below 50,000 ng/mL. ^b^Panel B insert shows in detail changes from baseline to week 4 for patients with baseline levels of aspartate aminotransferase below 300 U/L. ^c^Panel D insert shows in detail changes from baseline to week 4 for patients with baseline levels of LDH below 2500 U/L. LDH, lactate dehydrogenase.

During long-term follow-up, 13 patients did not have MAS episodes. One patient had a MAS episode 11 months after stopping emapalumab, when emapalumab was undetectable in serum. At the end of the long-term follow-up, 10 patients met the criteria of MAS remission. Four did not meet these criteria because of either mild abnormalities in one laboratory value (n=1), MAS activity at 1.5 cm (n=1), missing data (n=1) or absence of data as the patient missed the 12 month visit (Table S4).

Tapering of glucocorticoids occurred rapidly after emapalumab initiation. During the week preceding emapalumab, the median average daily dose was 15.7 mg/kg/day prednisone-equivalent, 2.3 mg/kg during week 2 and 0.56 mg/kg during week 8 (Fig. 4). At the end of the long-term follow-up, five patients were not receiving glucocorticoids and six were receiving <0.3 mg/kg/day prednisone-equivalent. Two patients were receiving doses between 1 and 2 mg/kg: the above-mentioned patient with a MAS episode and one patient with lung disease associated with sJIA. Data on glucocorticoid dose for one patient at the last follow-up visit are not available, as the patient missed the visit.

**Figure 4.**
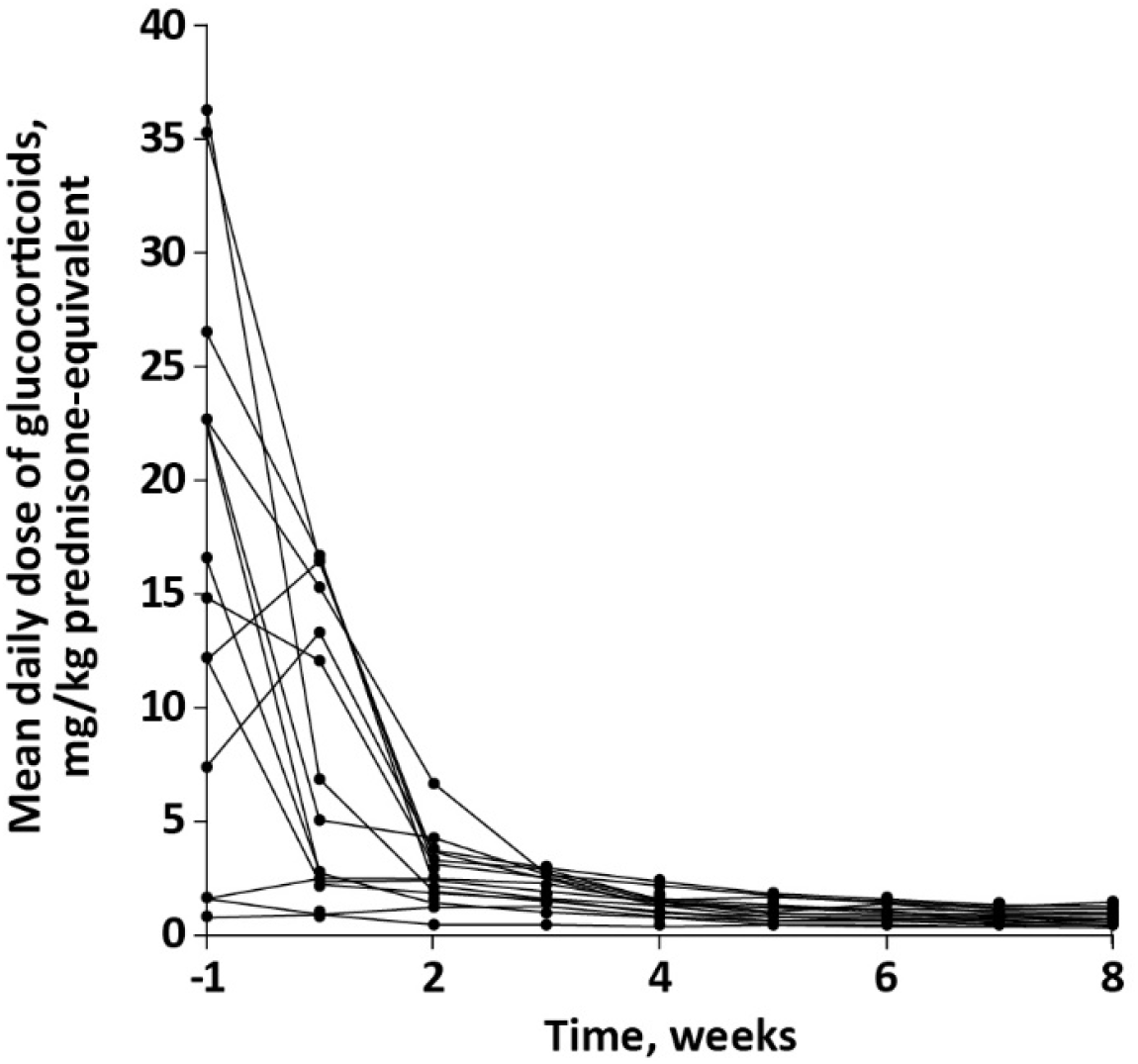
Changes in glucocorticoid dose^a^ over time. ^a^ The dose of glucocorticoids is shown as average daily dose in each week of the study, starting from the week preceding emapalumab treatment (week -1). The dose is shown in mg/kg of prednisone equivalent.

Eight patients were receiving ciclosporin at baseline. Ciclosporin was discontinued in two patients early after initiation of emapalumab (day 4 and 10) and in four additional patients during long-term follow-up.

During the trial, anakinra was continued in four patients at a dose of ≤4 mg/kg, the standard dose for sJIA/AOSD treatment, and in one patient at a dose of 7.5 mg/kg. These patients did not present with flares of the underlying sJIA/AOSD. During the trial, while MAS was improving, six flares of sJIA/AOSD were observed: three in the three patients who discontinued anakinra before emapalumab initiation and three in three patients who had not previously received and were not receiving anakinra. During long-term follow-up, four sJIA/AOSD flares occurred in four patients. sJIA/AOSD flares and the background treatment at time of the flares are described in Table S5.

### Safety

No deaths were reported during the trial and the long-term follow-up. During the trial, after initiation of emapalumab, 88 AEs were reported in 13 patients (Table 3 and Table S6). All events were mild or moderate in intensity except two (one cardiopulmonary failure, and one neutropenia; neither related to emapalumab). The most frequently reported AEs were infections and positive tests for infectious agents in the absence of clinical symptoms. All infectious events were of viral origin. No bacterial or opportunistic infections were reported. Six viral events in three patients (two infections and four positive tests) were reported as related to emapalumab. One cytomegalovirus (CMV) reactivation was reported as serious. In total, there were five CMV events (three reactivations, one infection and one positive test with no symptoms). All viral events resolved spontaneously or with standard treatment. Two infusion-related reactions (pruritic rash), not reported as severe, occurred during a total of 128 infusions. The rate of AEs and of infectious events was not increased during concomitant treatment with anakinra and emapalumab compared with treatment with emapalumab alone (Table 4).

**Table 3.**
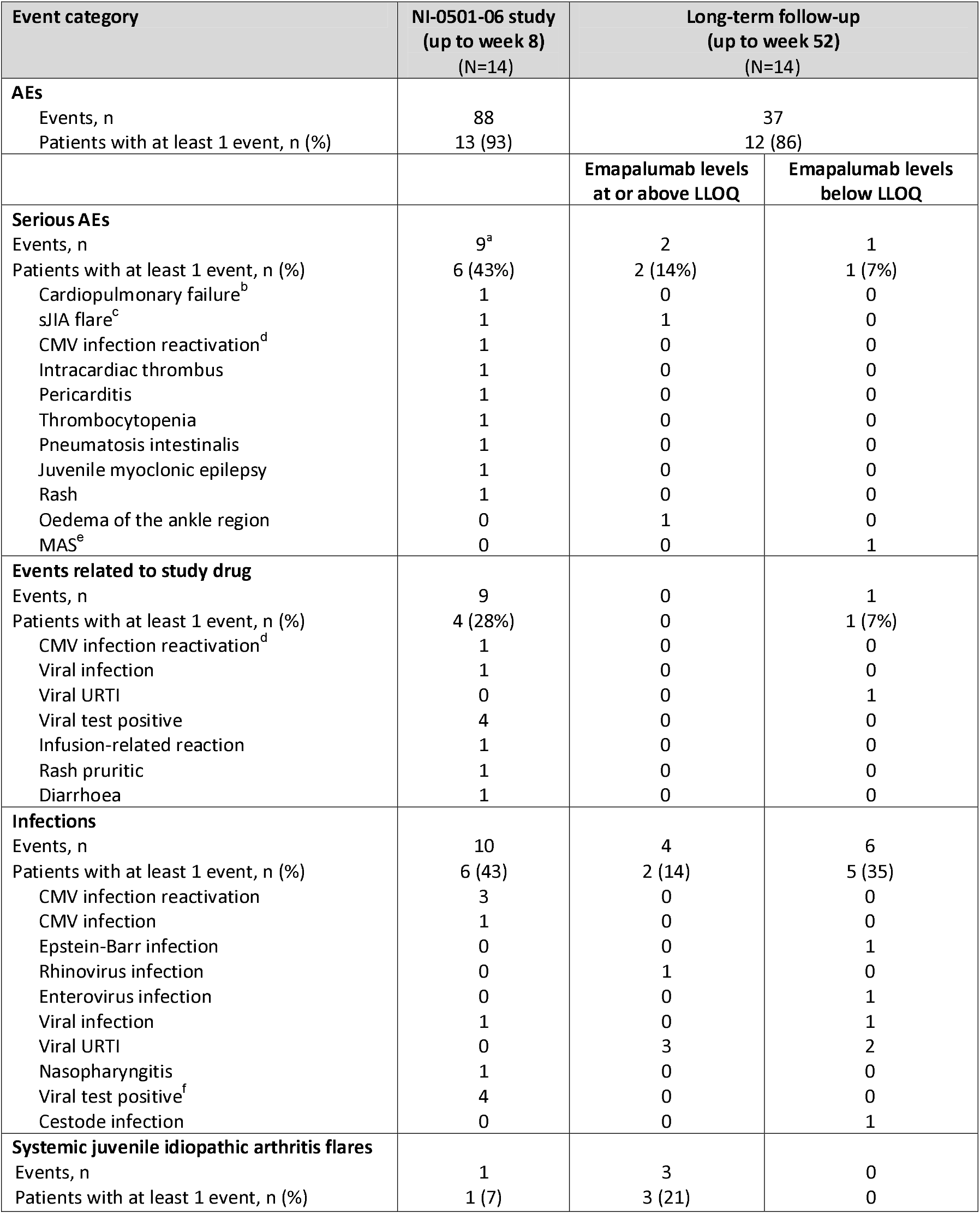
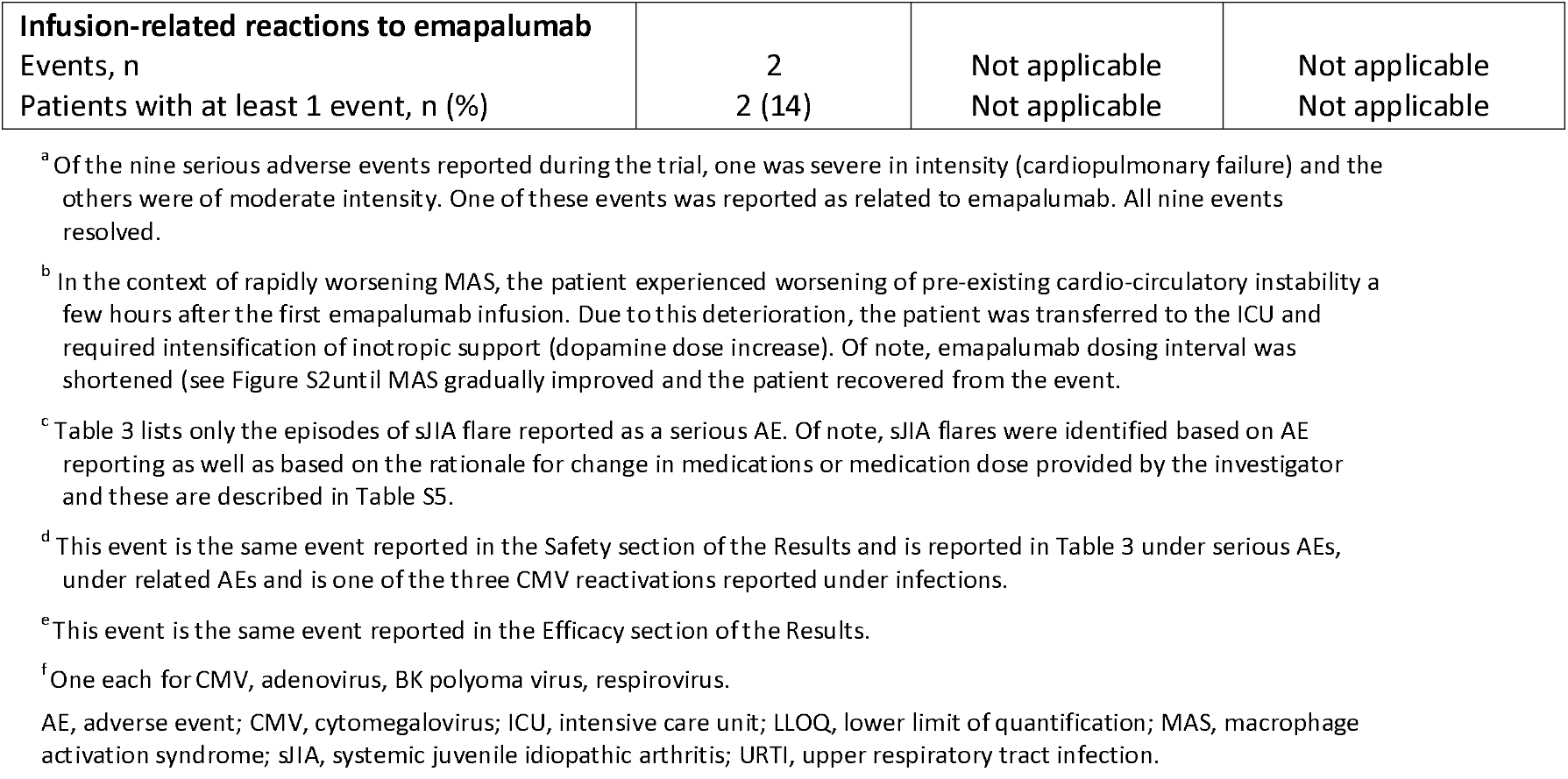
AEs during the treatment with emapalumab and during the long-term-follow-up.

**Table 4.**
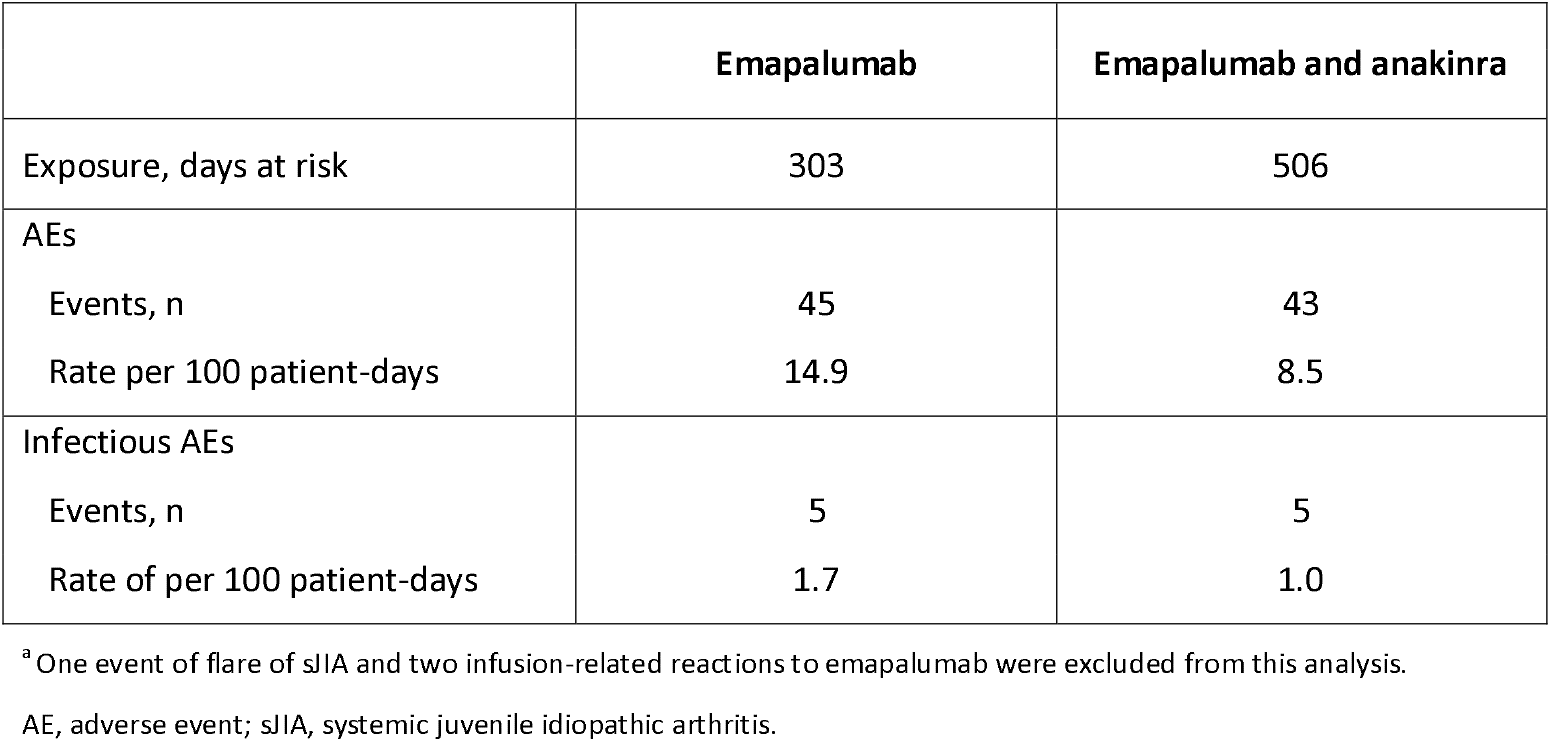
Rate of AEs^a^ and infectious AEs during the trial while exposed to emapalumab alone or emapalumab and anakinra.

During long-term follow-up, 37 AEs were reported in 12 patients. Three were serious: one flare of sJIA (see Table S5), one oedema of the ankle and one MAS episode (that, as mentioned above, occurred 11 months after stopping emapalumab, when emapalumab was undetectable). Ten infectious events occurred in seven patients; four when emapalumab levels were measurable, and six after emapalumab levels became undetectable. All reported infections were viral, except for one cestode infection.

## DISCUSSION

We show that neutralisation of IFNγ with emapalumab was efficacious in inducing MAS remission in patients with MAS secondary to sJIA/AOSD who failed standard of care with high-dose glucocorticoids.

The dosing regimen of emapalumab was chosen based on the data gathered in primary HLH patients on a) the production rate of IFNγ (through the assessment of total IFNγ), b) the rapid clearance of emapalumab consequent to target-mediated drug disposition in the presence of high IFNγ production and c) the concentration of emapalumab required to neutralise high IFNγ levels. Through modelling and simulation, on the basis of the high levels of CXCL9 present in MAS patients, it was possible to predict the dose of emapalumab required to achieve IFNγ neutralisation in this context. A dosing regimen with an initial dose of 6 mg/kg, followed by 3 mg/kg every 3 days maintenance doses, was selected to achieve rapid efficacy as MAS usually has acute onset and may worsen rapidly, becoming life-threatening. In the majority of patients, the chosen regimen rapidly achieved IFNγ neutralisation, shown by prompt decrease in serum CXCL9 levels and prompt clinical and laboratory response. Shortening of the dosing interval might be considered in patients with unsatisfactory response. Indeed, in two patients with initial unsatisfactory response, shortening of the dosing interval led to achievement of response. In these patients, we found high IFNγ production and low emapalumab concentrations due to rapid drug clearance consistent with target-mediated drug disposition.

The patients recruited in this study had markedly elevated levels of CXCL9, reflecting high IFNγ activity. This finding is consistent with previous observations in patients with MAS.[16,18] In contrast with primary HLH patients treated with emapalumab,[27] we did not find a correlation of CXCL9 levels at baseline with total IFNγ levels at day 3, that, as previously mentioned, reflect IFNγ production. Therefore, in MAS, the pathogenic role of IFNγ may be due not only to increased IFNγ production, but also to increased sensitivity to IFNγ. Indeed, monocytes from patients with MAS have increased responsiveness to IFNγ, possibly through increased expression of TRIM-8, which potentiates response to IFNγ.[29,30]

In this trial, the first prospectively investigating a treatment for MAS, we used a novel clinically meaningful efficacy measure, namely MAS remission, that combines resolution of clinical signs and symptoms and resolution of the abnormalities in MAS laboratory parameters. After emapalumab initiation, MAS improved rapidly with median time to MAS remission of 25 days. This improvement occurred while background glucocorticoids were rapidly tapered: during the second week of emapalumab treatment, the median glucocorticoid dose was already 85% lower compared with that administered during the week preceding initiation of emapalumab. All patients benefited from emapalumab even when the strict criteria for MAS remission were only partially achieved. In the patients who did not reach MAS remission, the physician assessment of disease activity was ≤1 and MAS remission was not achieved because of minor abnormalities in only one of the laboratory parameters of MAS.

Elevated production of IFNγ is a feature of animal models of primary HLH, as well as of secondary HLH, including infection-associated HLH and MAS.[13] In these models, when the effect of IFNγ neutralisation was tested, benefit was always observed with prevention of death and/or improvement of disease. The results of this trial, together with the efficacy of emapalumab demonstrated in primary HLH,[27] and the anecdotal cases of patients with different forms of secondary HLH successfully treated with emapalumab,[31-37] suggest that, in humans, IFNγ is an important driver of MAS/HLH, independently of the trigger or the underlying predisposing condition.

Notably, some inflammatory flares of the underlying sJIA/AOSD were observed, suggesting that, in the context of sJIA/AOSD, increased IFNγ activity is a feature of the hyperinflammation of MAS, but not of the autoinflammation that maintains sJIA. From a clinical perspective, we observed six flares of the underlying sJIA/AOSD in six out of the nine patients who were not receiving anakinra for the treatment of sJIA while receiving emapalumab. In contrast, no sJIA flares were observed in the five patients who continued anakinra after emapalumab initiation. Importantly, during the trial no increase in AEs, and, in particular, of infectious AEs was observed in patients concomitantly exposed to emapalumab and anakinra. Therefore, continuation of anakinra for the underlying sJIA/AOSD, at the doses conventionally used and allowed in this trial, is an option to be considered.[38-40]

The potential contribution of the concomitant anakinra to the achievement of MAS remission with emapalumab is to be considered negligible. Only one patient continued anakinra at a dose >4 mg/kg/day, a so called higher dose that has been reported as potentially efficacious in MAS.[10,14,40] Of note, several patients recruited to our study received anakinra before treatment with emapalumab and, in these patients, anakinra, even at doses higher than 4 mg/kg/day, did not prevent or improve MAS. Furthermore, of the three patients in the trial who did not meet the criteria for MAS remission at week 8, two were receiving concomitant anakinra. We did not explore combining higher dose anakinra with emapalumab.

IFNγ neutralisation might increase predisposition to selected infections, as inferred based on data in humans with defective IFNγ activity, i.e., subjects carrying autoantibodies to IFNγ or with IFNγ receptor deficiency.[42] While herpes zoster infections are common in individuals with defective IFNγ activity, no cases were reported in this trial. It should be noted that, per-protocol, patients received prophylaxis with acyclovir. We did not observe any infections from typical or atypical mycobacteria, *Histoplasma capsulatum, Shigella, Salmonella, Campylobacter* or *Leishmania*, known to occur more frequently in subjects with defective IFNγ activity. Noteworthy, patients with infections from these agents were excluded from the trial. No bacterial or opportunistic infections were reported. Viral infections or positive viral tests were reported in six patients during the trial and in two during follow-up while emapalumab was detectable. All events resolved spontaneously or with standard treatment. Of note, screening for Epstein-Barr virus, CMV and adenovirus every 2 weeks was required by protocol during the trial. Despite the fact that CMV is not reported at increased frequency in humans with defective IFNγ activity, [42] it should be noted that five events in five patients were related to CMV, with four reported as reactivations or infections and one as test positive. Whether concomitant prolonged immunosuppression with high-dose glucocorticoids in critically ill patients might have contributed to this observation remains to be established. Despite CMV infections not being reported in patients defective IFNγ activity in humans, [42] based on our data, patients with MAS receiving emapalumab should be screened for the presence of CMV.

In conclusion, the results of this trial demonstrate that IFNγ is an important driver of MAS secondary to sJIA/AOSD and that its neutralisation with emapalumab leads to remission of MAS in patients who failed high-dose glucocorticoids. Attention should be paid to viral infections, particularly to CMV, and periodic screening for viral infections should be performed.

## Data Availability

Data are available on reasonable request. The data sharing policy of Sobi is available at the following site: https://www.sobi.com/en/policies. As noted on that site, requests for access to the study data can be submitted to.

## Ethics statements

### Patient consent for publication

Not applicable.

### Ethics approval

Approval was obtained from relevant regional institutional review boards or independent ethics committees. All patients or their legal representatives provided written informed consent before taking part.

## Acknowledgements

The authors wish to acknowledge the contribution of the study participants and of their families. Medical writing assistance was provided by Blair Hesp PhD CMPP of Kainic Medical Communications Ltd. (Dunedin, New Zealand), which was funded by Sobi (Basel, Switzerland). PQ thanks Ms Thi An Vu (Clinical Research Unit, Necker Hospital, Paris, France) for her support. PB acknowledges the conduct of the trial at Great Ormond Street Hospital NHS Foundation Trust and UCL Great Ormond Street Institute of Child Health being supported by the NIHR Great Ormond Street Hospital Biomedical Research Centre. AG acknowledges PRCSG Clinical Research Associates at Cincinnati Children’s Hospital.

## Footnotes

### Presented at

Part of the content in this manuscript has previously been presented at the American College of Rheumatology Convergence 2021, the 37th Annual Meeting of the Histiocyte Society, the 63rd American Society of Hematology Annual Meeting and Exposition, the European Alliance for Associations of Rheumatology 2022 Congress, the American Society of Pediatric Hematology and Oncology Conference 2022 and the Paediatric Rheumatology European Society 2022 Congress.

### Contributors

FDB, AG, CdM, MB and PJ provided substantial contributions to the conception of the work. All authors substantially contributed to the acquisition, analysis or interpretation of data for the manuscript. All authors participated in drafting, revising and critically reviewing the manuscript for important intellectual content. All authors approved final version of this manuscript to be published and agree to be accountable for all aspects of the work in ensuring that questions related to the accuracy or integrity of any part of the work are appropriately investigated and resolved.

### Patient and public involvement

The research program and the design of the study were presented and discussed at the 2016 and 2017 meeting of the sJIA Foundation in Washington, DC, USA. Information on the study to patients and families were published on the sJIA Foundation website. Dissemination of the data gathered from the study is planned to be discussed also with sJIA Foundation.

### Provenance and peer review

Not commissioned; externally peer reviewed.

### Supplemental material

This content has been supplied by the author(s). It has not been vetted by BMJ Publishing Group Limited (BMJ) and may not have been peer-reviewed. Any opinions or recommendations discussed are solely those of the author(s) and are not endorsed by BMJ. BMJ disclaims all liability and responsibility arising from any reliance placed on the content. Where the content includes any translated material, BMJ does not warrant the accuracy and reliability of the translations (including but not limited to local regulations, clinical guidelines, terminology, drug names and drug dosages), and is not responsible for any error and/or omissions arising from translation and adaptation or otherwise.

## REFERENCES

1. Grom AA, Horne A, De Benedetti F. Macrophage activation syndrome in the era of biologic therapy. Nat Rev Rheumatol 2016;12:259–68.

2. Efthimiou P, Kontzias A, Hur P, et al. Adult-onset Still’s disease in focus: Clinical manifestations, diagnosis, treatment, and unmet needs in the era of targeted therapies. Semin Arthritis Rheum 2021;51:858–874.

3. Jamilloux Y, Gerfaud-Valentin M, Martinon F, et al. Pathogenesis of adult-onset Still’s disease: new insights from the juvenile counterpart. Immunol Res 2015;61:53–62.

4. Minoia F, Davì S, Horne A, et al. Clinical features, treatment, and outcome of macrophage activation syndrome complicating systemic juvenile idiopathic arthritis: a multinational, multicenter study of 362 patients. Arthritis Rheumatol 2014;66:3160–9.

5. Nigrovic PA, De Benedetti F, Kimura Y, et al. The elephant in the room: diagnostic/classification criteria for systemic juvenile idiopathic arthritis and adult-onset Still’s disease. Pediatr Rheumatol 2022;In press.

6. De Matteis A, Colucci M, Rossi MN, et al. Expansion of CD4dimCD8+T cells characterizes macrophage activation syndrome and other secondary HLH. Blood 2022;140:262–73.

7. Chaturvedi V, Marsh RA, Zoref-Lorenz A, et al. T-cell activation profiles distinguish hemophagocytic lymphohistiocytosis and early sepsis. Blood 2021;137:2337–46.

8. Erkens R, Esteban Y, Towe C, et al. Pathogenesis and treatment of refractory disease courses in systemic juvenile idiopathic arthritis: Refractory arthritis, recurrent macrophage activation syndrome and chronic lung disease. Rheum Dis Clin North Am 2021;47:585–606.

9. Horne A, von Bahr Greenwood T, Chiang SCC, et al. Efficacy of moderately dosed etoposide in macrophage activation syndrome-hemophagocytic lymphohistiocytosis. J Rheumatol 2021;48:1596–602.

10. Eloseily EM, Weiser P, Crayne CB, et al. Benefit of anakinra in treating pediatric secondary hemophagocytic lmphohistiocytosis. Arthritis Rheumatol 2020;72:326–34.

11. Boom V, Antón J, Lahdenne P, et al. Evidence-based diagnosis and treatment of macrophage activation syndrome in systemic juvenile idiopathic arthritis. Pediatr Rheumatol Online J 2015;13:55.

12. Verweyen EL, Schulert GS. Interfering with interferons: targeting the JAK-STAT pathway in complications of systemic juvenile idiopathic arthritis (SJIA). Rheumatology (Oxford) 2022;61:926–35.

13. De Benedetti F, Prencipe G, Bracaglia C, et al. Targeting interferon-γ in hyperinflammation: opportunities and challenges. Nat Rev Rheumatol 2021;17:678–91.

14. Girard-Guyonvarc’h C, Palomo J, Martin P, et al. Unopposed IL-18 signaling leads to severe TLR9-induced macrophage activation syndrome in mice. Blood 2018;131:1430–41.

15. Prencipe G, Caiello I, Pascarella A, et al. Neutralization of IFN-γ reverts clinical and laboratory features in a mouse model of macrophage activation syndrome. J Allergy Clin Immunol 2018;141:1439–49.

16. Weiss ES, Girard-Guyonvarc’h C, Holzinger D, et al. Interleukin-18 diagnostically distinguishes and pathogenically promotes human and murine macrophage activation syndrome. Blood 2018;131:1442–55.

17. Groom JR, Luster AD. CXCR3 ligands: redundant, collaborative and antagonistic functions. Immunol Cell Biol 2011;89:207–15.

18. Bracaglia C, de Graaf K, Pires Marafon D, et al. Elevated circulating levels of interferon-γ and interferon-γ-induced chemokines characterise patients with macrophage activation syndrome complicating systemic juvenile idiopathic arthritis. Ann Rheum Dis 2017;76:166–72.

19. Mizuta M, Shimizu M, Inoue N, et al. Clinical significance of serum CXCL9 levels as a biomarker for systemic juvenile idiopathic arthritis associated macrophage activation syndrome. Cytokine 2019;119:182–7.

20. Rodriguez-Smith JJ, Verweyen EL, Clay GM, et al. Inflammatory biomarkers in COVID-19-associated multisystem inflammatory syndrome in children, Kawasaki disease, and macrophage activation syndrome: a cohort study. Lancet Rheumatol 2021;3:e574–84.

21. Takakura M, Shimizu M, Irabu H, et al. Comparison of serum biomarkers for the diagnosis of macrophage activation syndrome complicating systemic juvenile idiopathic arthritis. Clin Immunol 2019;208:108252.

22. Han JH, Suh CH, Jung JY, et al. Elevated circulating levels of the interferon-γ-induced chemokines are associated with disease activity and cutaneous manifestations in adult-onset Still’s disease. Sci Rep 2017;7:46652.

23. Petty RE, Southwood TR, Manners P, et al. International League of Associations for Rheumatology classification of juvenile idiopathic arthritis: second revision, Edmonton, 2001. J Rheumatol 2004;31:390–2.

24. Yamaguchi M, Ohta A, Tsunematsu T, et al. Preliminary criteria for classification of adult Still’s disease. J Rheumatol 1992;19(3):424-30. (In eng).

25. DeWitt EM, Kimura Y, Beukelman T, et al. Consensus treatment plans for new-onset systemic juvenile idiopathic arthritis. Arthritis Care Res (Hoboken) 2012;64:1001–10.

26. Ravelli A, Minoia F, Davì S, et al. 2016 Classification criteria for macrophage activation syndrome complicating systemic juvenile idiopathic arthritis: A European League Against Rheumatism/American College of Rheumatology/Paediatric Rheumatology International Trials Organisation collaborative initiative. Arthritis Rheumatol 2016;68:566–76.

27. Locatelli F, Jordan MB, Allen C, et al. Emapalumab in children with primary hemophagocytic lymphohistiocytosis. N Engl J Med 2020;382:1811–22.

28. Jacqmin P, Laveille C, Snoeck E, et al. Emapalumab in primary haemophagocytic lymphohistiocytosis and the pathogenic role of interferon gamma: A pharmacometric model-based approach. Br J Clin Pharmacol 2022;88:2128–39.

29. Pascarella A, Bracaglia C, Caiello I, et al. Monocytes from patients with macrophage activation syndrome and secondary hemophagocytic lymphohistiocytosis are hyperresponsive to interferon gamma. Front Immunol 2021;12:663329.

30. Schulert GS, Pickering AV, Do T, et al. Monocyte and bone marrow macrophage transcriptional phenotypes in systemic juvenile idiopathic arthritis reveal TRIM8 as a mediator of IFN-γ hyper-responsiveness and risk for macrophage activation syndrome. Ann Rheum Dis 2021;80:617–25.

31. Lounder DT, Bin Q, de Min C, et al. Treatment of refractory hemophagocytic lymphohistiocytosis with emapalumab despite severe concurrent infections. Blood Adv 2019;3:47–50.

32. McNerney KO, DiNofia AM, Teachey DT, et al. Potential role of IFNγ inhibition in refractory cytokine release syndrome associated with CAR T-cell therapy. Blood Cancer Discov 2022;3:90–4.

33. Tucci F, Gallo V, Barzaghi F, et al. Emapalumab treatment in an ADA-SCID patient with refractory hemophagocytic lymphohistiocytosis-related graft failure and disseminated bacillus Calmette-Guérin infection. Haematologica 2021;106:641–6.

34. Lam MT, Coppola S, Krumbach OHF, et al. A novel disorder involving dyshematopoiesis, inflammation, and HLH due to aberrant CDC42 function. J Exp Med 2019;216:2778–99.

35. Triebwasser MP, Barrett DM, Bassiri H, et al. Combined use of emapalumab and ruxolitinib in a patient with refractory hemophagocytic lymphohistiocytosis was safe and effective. Pediatr Blood Cancer 202;68:e29026.

36. Shamriz O, Kumar D, Shim J, et al. T cell-Epstein-Barr virus-associated hemophagocytic lymphohistiocytosis (HLH) occurs in non-Asians and is associated with a T cell activation state that is comparable to primary HLH. J Clin Immunol 2021;41:1582–96.

37. Gabr JB, Liu E, Mian S, et al. Successful treatment of secondary macrophage activation syndrome with emapalumab in a patient with newly diagnosed adult-onset Still’s disease: case report and review of the literature. Ann Transl Med 2020;8:887.

38. Urien S, Bardin C, Bader-Meunier B, et al. Anakinra pharmacokinetics in children and adolescents with systemic-onset juvenile idiopathic arthritis and autoinflammatory syndromes. BMC Pharmacol Toxicol 2013;14:40.

39. Vastert SJ, Jamilloux Y, Quartier P, et al. Anakinra in children and adults with Still’s disease. Rheumatology (Oxford) 2019;58(Suppl 6):vi9–vi22.

40. KINERET (anakinra). Summary of product characteristics. https://www.ema.europa.eu/en/documents/product-information/kineret-epar-product-information_en.pdf

41. Mehta P, Cron RQ, Hartwell J, et al. Silencing the cytokine storm: the use of intravenous anakinra in haemophagocytic lymphohistiocytosis or macrophage activation syndrome. Lancet Rheumatol 2020;2:e358–67.

42. Shih HP, Ding JY, Yeh CF, et al. Anti-interferon-γ autoantibody-associated immunodeficiency. Curr Opin Immunol 2021;72:206–14.

