## Supplementary Materials for "Efficacy and safety of emapalumab in macrophage activation syndrome"

**Supplementary Information**

**Appendix 1. Total interferon-γ, CXCL9, and soluble interleukin-2 receptor assays**

Validated MesoScale Discovery (Rockville, MD, USA)-based immunoassays were used for the measurement of total interferon-γ (IFNγ), soluble interleukin-2 receptor and CXCL9. The respective ranges of quantification relative to the recombinant protein were: interferon-γ, 50 to 50,000 pg/mL; soluble interleukin-2 receptor (sIL-2R), 50 to 50,000 pg/mL; CXCL9, 80 to 50,000 pg/mL. Samples containing biomarker concentrations above these levels underwent dilution to achieve the working range of quantification, with the final result corrected for the sample dilution used. Runs failing to meet standard and/or quality controls (QCs) acceptance criteria were repeated.

Total IFNγ

Calibration standard (recombinant human IFNγ, R&D Systems, Minneapolis, MN, USA) ranging from 20 to 100,000 pg/mL was prepared in human pool serum (at least 10 individual human sera, Seralab/BioIVT, West Sussex, UK) and incubated 1h at 22°C in the presence of 50 ug/mL final of emapalumab to mimic clinical samples. Streptavidin Gold plates (MesoScale Discovery, Rockville, MD, USA) were pre-washed three times with phosphate-buffered saline-Tween 0.05% (wash buffer). Plates were coated with 25 µL of biotinylated mouse monoclonal antibody anti-human IFNγ clone 1-D1K (Mabtech, Nacka Strand, Sweden) at 0.50 μg/mL in phosphate-buffered saline-bovine serum albumin (PBS-BSA) 1%-Tween 0.05% and incubated 1h at 22°C with agitation at 600 rpm. Samples were tested neat or diluted 1:50 in pool serum. Three QC samples were made of recombinant human IFNγ diluted in human pool serum respectively at 160, 2000, and 40,000 pg/mL with a final concentration of emapalumab of 50 ug/mL. Standard samples and QC samples were diluted at the minimum required dilution (1:3) in PBS-BSA 1%-Tween 0.05% with a final concentration of emapalumab of 50 ug/mL and incubated 1 h at 22°C. Streptavidin Gold plates were washed three times and 25 µL of standard, samples and QC samples were added per well, in duplicate. After 1h incubation at 22°C with agitation at 600 rpm, plates were washed three times and 25 µL of Sulfo-TAG® (MesoScale Discovery, Rockville, MD, USA)-labelled mouse monoclonal antibody anti-human IFNγ clone 1-D1K (Mabtech, Nacka Strand, Sweden) at 0.60 μg/mL in PBS-BSA 1%-Tween 0.05% were added per well. After 1 h incubation at 22°C with agitation at 600 rpm, plates were washed three times and 150 μL of MSD read buffer T 2X (MesoScale Discovery, Rockville, MD, USA) were added per well and the plates were read within 4 to 10 min on the MesoScale Discovery Meso Sector® S600. The raw data were analysed using the MesoScale Discovery Discovery Workbench 4.0 software.

Soluble interleukin-2 receptor

Streptavidin Gold plates (MesoScale Discovery, Rockville, MD USA) were pre-washed three times with PBS-Tween 0.05% (wash buffer). Plates were coated with 25 µL of biotinylated polyclonal goat immunoglobulin G anti-human CD25/interleukin-2 receptor α (R&D systems, Minneapolis, MN, USA) at 0.50 μg/mL in PBS-BSA 1% and incubated 1h at 22°C with agitation at 600 rpm. Calibration standard (recombinant human interleukin receptor 2α, Reprokine, St Petersburg, FL, USA) ranging from 20 to 100,000 pg/mL was prepared in foetal bovine serum (Sigma-Aldrich, Saint Louis, MO, USA). Samples were tested neat or diluted 1:2, 1:5, 1:10, 1:20 1:50 in foetal bovine serum. Three quality control samples were made of recombinant human sIL-2Rα spiked in foetal bovine serum at respectively 35,000 pg/mL, 1800 pg/mL and 150 pg/mL. Two MatrixQCs were made in human pool serum, one containing endogenous sIL-2Rα only, the other containing endogenous sIL-2Rα plus spiked recombinant sIL-2Rα. Standard, samples, QC samples and MatrixQCs were diluted at the minimum required dilution (1:10) in PBS-BSA 1%. Streptavidin Gold plates were washed three times and 25 µL of standard, samples, QC samples and MatrixQCs were added per well, in duplicate. After 1 h incubation at 22°C with agitation at 600 rpm, plates were washed three times and 25 µL of monoclonal mouse immunoglobulin G1 clone 24204 anti-human CD25/interleukin 2 receptor α (R&D systems, Minneapolis, MN, USA) labelled with Sulfo-TAG® (MesoScale Discovery, Rockville, MD, USA) at 0.80 or 1.20 μg/mL in PBS-BSA 1% were added per well. After 1 h incubation at 22°C with agitation at 600 rpm, plates were washed three times and 150 μL of MesoScale Discovery read buffer T 2X (MesoScale Discovery, Rockville, MD, USA) were added per well and the plates were read within 1 to 10 min on the MesoScale Discovery Meso Sector® S600. The raw data were analysed using the MesoScale Discovery Discovery Workbench 4.0 software.

CXCL9

Streptavidin Gold plates (MesoScale Discovery, Rockville, MD, USA) were pre-washed three times with PBS-Tween 0.05% (wash buffer). Plates were coated with 25 µL of mouse anti-human MIG biotinylated antibody (MesoScale Discovery, Rockville, MD, USA) at 0.25 μg/mL in PBS-BSA 1% and incubated 1 h at 22°C with agitation at 600 rpm. Calibration standard (recombinant human CXCL9, R&D systems, Minneapolis, MN, USA) ranging from 30 to 100,000 pg/mL was prepared in foetal bovine serum (Sigma-Aldrich, Saint Louis, MO, USA). Samples were tested neat or diluted 1:2, 1:5, 1:10, 1:20 and 1:50 in foetal bovine serum. Three MatrixQCs were made of human pool serum containing endogenous CXCL9 only or containing endogenous plus spiked recombinant CXCL9. Standard, samples and MatrixQCs were diluted at the minimum required dilution (1:10) in PBS-BSA 1%. Streptavidin Gold plates were washed three times and 25 µL of standard, samples and QC samples were added per well, in duplicate. After 1 h incubation at 22°C with agitation at 600 rpm, plates were washed three times and 25 µL of goat anti-human MIG antibody labelled with Sulfo-TAG (MesoScale Discovery, Rockville, MD, USA) at 0.25X in PBS-BSA 1% were added per well. After 1 h incubation at 22°C with agitation at 600 rpm, plates were washed three times and reading was performed similarly as described above for sIL-2Rα.

**Appendix 2. Emapalumab assay**

A validated Gyrolab (Gyros Protein Technologies AB, Uppsala, Sweden)-based immunoassay was used for the measurements of emapalumab in patient’s serum. Quantification range was from 62.5 ng/mL to 8000 ng/mL. Samples containing emapalumab concentrations above these levels underwent dilution to achieve the working range of quantification, with the final result corrected for the sample dilution used. Runs failing to meet standard and/or quality control (QC) acceptance criteria were repeated.

Working calibration standards ranging from 31.25 to 16,000 ng/mL were freshly prepared in pooled human serum (at least 10 individuals, Seralab/BioIVT, West Sussex, UK) on the day of the assay. Two sets of QC samples thawed on the day of the assay were included on each compact disk (CD) and were comprised of pooled human serum, spiked with emapalumab at three concentration levels, 140 ng/mL (lower QC), 850 ng/mL (medium QC) and 6000 ng/mL (high QC). Two additional QC samples at 250,000 and 12,000 ng/mL were added to runs with clinical samples having a corresponding dilution to assess the validity of the 1:60 dilution. These QC samples were prepared by spiking reference standard into human pool serum and were diluted 1:60 in pooled human serum prior to dilution to the minimum required dilution.

Clinical samples were tested preferentially at the minimum required dilution or diluted 1:60 in addition to the minimum required dilution using pooled human serum, based on the amount of emapalumab expected to be present in the sample. QC samples and samples were diluted to a minimum required dilution of 1:3 in buffer Rexxip H (Gyros Protein Technologies AB, Uppsala, Sweden). Only one replicate of standards, QC samples and samples were prepared. The replicate was analysed twice by the Gyrolab Workstation on the CD to create the duplicate.

The CD contains micro-affinity columns which are nanostructures packed with streptavidin coated beads. The diluted samples, standard and QC samples as well as the reagents (capture and detection antibodies, wash solution) are prepared manually and loaded in a Gyrolab xP instrument via a polymerase chain reaction plate, which performs the pipetting, incubation, and wash steps of the immunoassay according to a set program where the plate contents are delivered to reservoirs in the CD and the spinning action of the CD causes them to flow through the column. A biotinylated monoclonal anti-idiotypic antibody specific to emapalumab diluted at 52.38 μg/mL with biotinylated bovine serum albumin at 22.73 μg/mL in phosphate-buffered saline-Tween 0.01% was added to the columns, allowing the biotin to bind to the streptavidin beads. The columns were washed, and samples, standards or controls added. After an additional wash the monoclonal anti-idiotypic detection antibody specific to emapalumab coupled with a fluorescent label diluted at 16.67 nM in Rexxip F (Gyros Protein Technologies AB, Uppsala, Sweden) was added. After a final wash step, the Gyrolab instrument measured the fluorescence in all of the columns, simultaneously. The raw data were analysed with Gyrolab Evaluator software.

**Figure S1. Design of the NI-0501-06 study and of the NI-0501-05 long-term follow-up study**

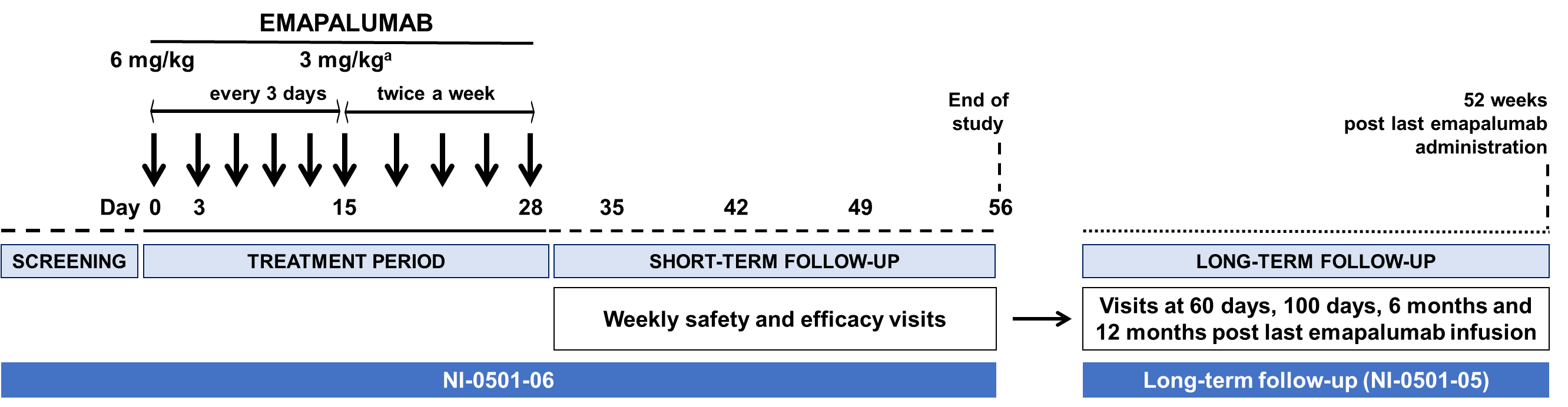
^a^ Treatment with emapalumab could be shortened upon achievement of macrophage activation syndrome remission as per investigator’s assessment, but not prior to the third dose; frequency between infusions could be shortened or dose could be increased or treatment prolonged in the absence of satisfactory response as per investigator’s assessment.

**Figure S2.** **Concentration-time profiles of emapalumab (blue, left log axis), ferritin (dark red, left log axis), total IFNγ (orange, right log axis), sIL-2R (red, right log axis) and CXCL9 (green, right log axis) in two patients showing initial rapid drug clearance with high levels of total IFNγ and incomplete neutralisation of IFNγ activity, as demonstrated by CXCL9 levels and insufficient initial response, as shown by ferritin levels**

**Patient 2**

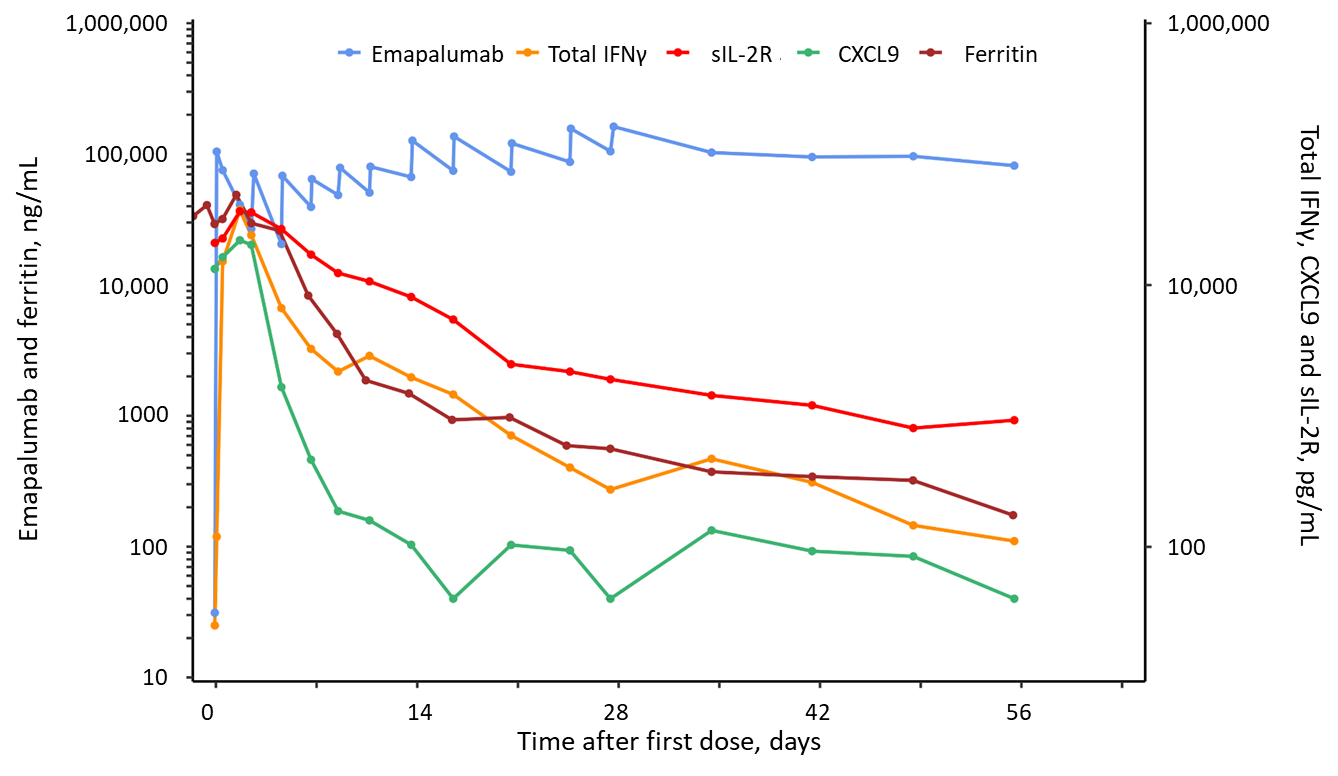

Patient 2 presented with rapidly worsening MAS leading to multiple organ failure and hypotensive shock not responding to multiple 1 g methylprednisolone pulses and required admission to intensive care unit. After start of emapalumab, CXCL9 levels, reflecting high IFNγ activity, and ferritin levels, indicating poor control of MAS, continued to increase. At the same time, the rapid decrease of emapalumab levels between infusions, together with high levels of total IFNγ (indicative of high IFNγ production) are consistent with the presence of target-mediated drug disposition. After shortening the dosing interval to 2 days (from day 5 to day 11), emapalumab levels started to increase and this was associated with neutralisation of IFNγ activity, as shown by decreased in CXCL9 levels and by a subsequent decrease in ferritin levels (and of all other MAS parameters [not shown]), reflecting progressive control of MAS activity.

**Patient 5**

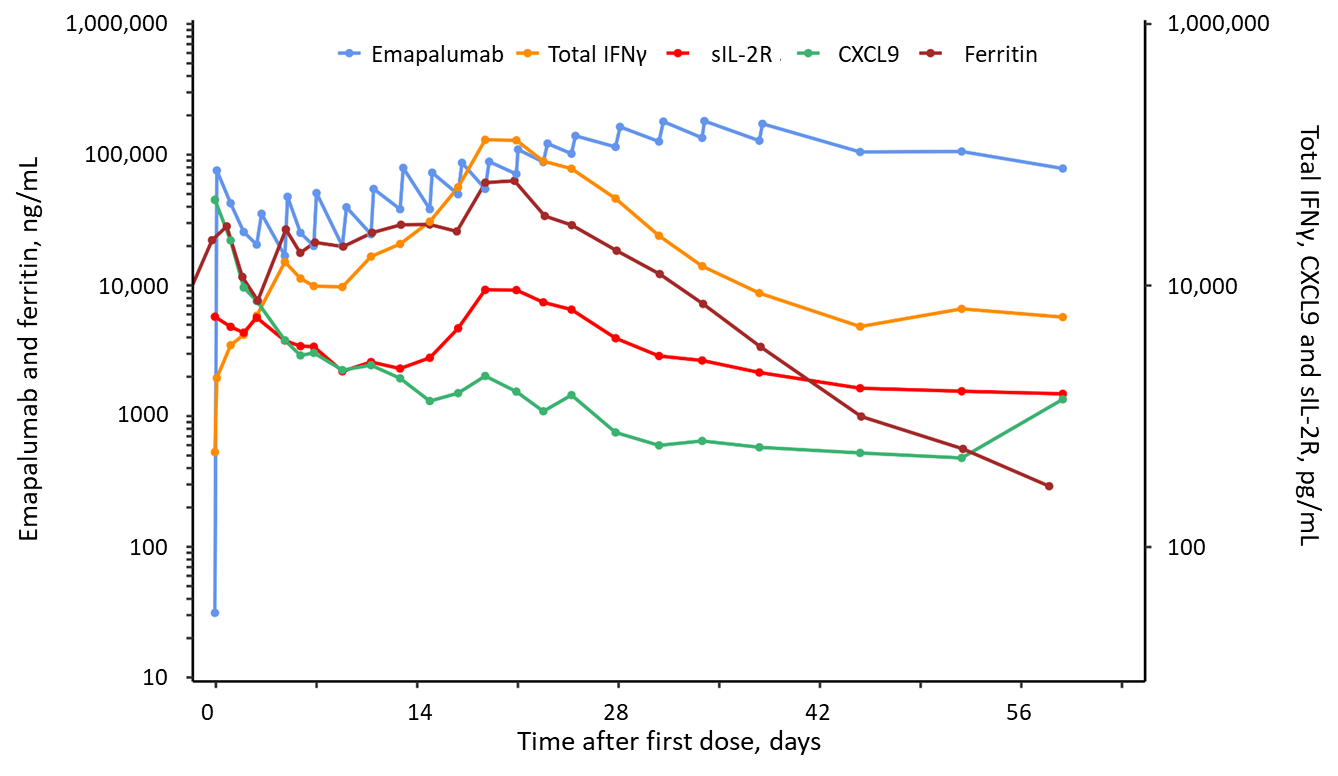

Patient 5 presented with chronic relapsing MAS having failed multiple treatments in the previous 2 months, including multiple pulses of glucocorticoids, anakinra and tocilizumab. After an initial response to emapalumab, with an evident decrease in ferritin levels, an increase in ferritin is observed, occurring at the time of CMV reactivation. The concomitant rapid decrease of emapalumab levels between infusions, together with persistently high levels of total IFNγ (indicative of high IFNγ production) are consistent with the presence of target mediated drug disposition. Sustained increase in levels of total IFNγ (indicative of high IFNγ production), of sIL-2R (reflecting T cell activation) and of ferritin (reflecting poor control of MAS activity) subsided after shortening the dosing interval to 2 days (from day 3 to day 25). Progressive control of MAS activity was achieved.

Abbreviations:

CMV, cytomegalovirus; IFNγ, interferon gamma; MAS, macrophage activation syndrome; sIL-2R, soluble interleukin-2 receptor

**Figure S3. Serum emapalumab levels during the NI-0501-06 study and the long-term follow-up study**

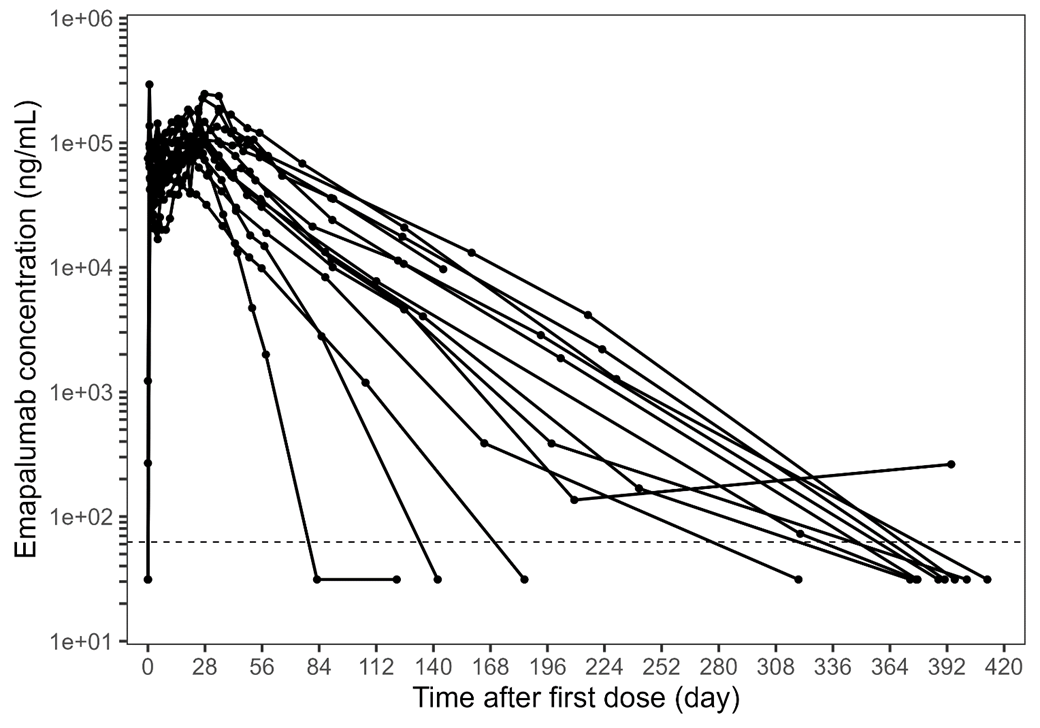

Median half-life during the long-term follow-up in the absence of interferon-γ production was 24 days, with a range from 6.1 to 32.4 days.

Dotted line represents the lower limit of quantification (62.5 ng/mL).

**Figure S4. Laboratory parameters of MAS, in addition to those shown in Fig 3, of individual patients depicting WBCs (A), fibrinogen (B), alanine aminotransferase (C) before, during and after emapalumab treatment**

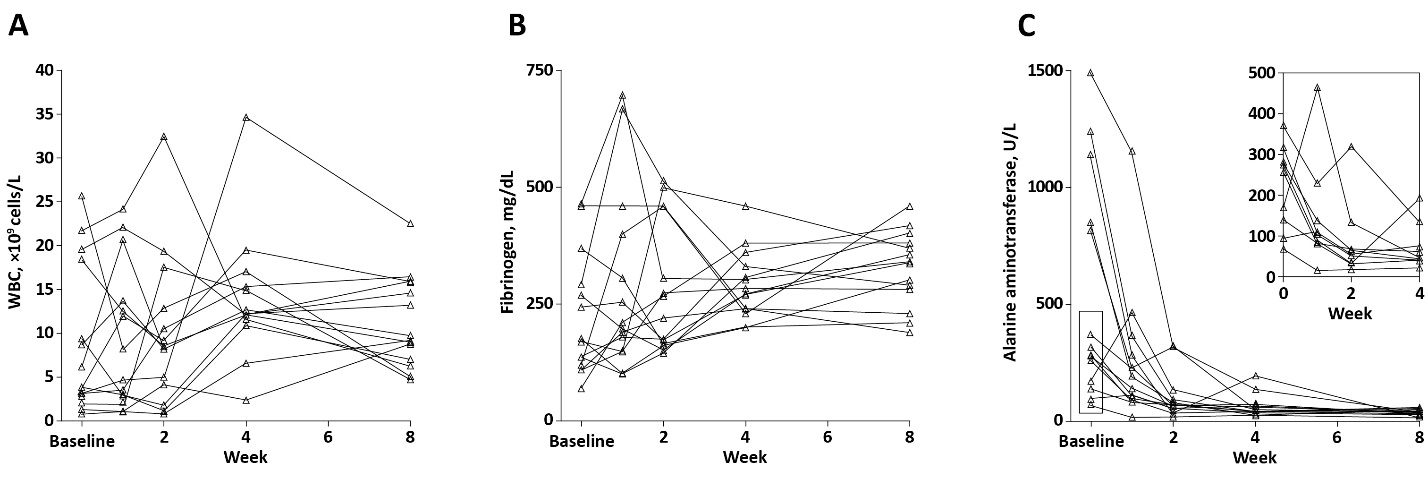

The insert in panel C shows in detail the change from baseline to week 4 in alanine aminotransferase for patients with baseline levels of alanine aminotransferase below 500 U/L.

Abbreviations:

WBC, white blood cells

**Table S1.** **Eligibility criteria (inclusion and exclusion) of the NI-0501-06 study**

**Inclusion criteria**

- Patients of both genders.
- For patients with sJIA: Confirmed sJIA diagnosis. For patients presenting with MAS in the context of the onset of sJIA, high presumption of sJIA sufficed for eligibility. For AOSD patients: confirmed AOSD diagnosis as per Yamaguchi criteria.
- A diagnosis of active MAS confirmed by the treating rheumatologist, having ascertained the following:

Febrile patient presenting with:

- Ferritin >684 ng/mL

and any 2 of:

- Platelet count ≤181 x10^9^ cells/L
- Aspartate aminotransferase levels >48 U/L
- Triglycerides >156 mg/dL
- Fibrinogen levels ≤360 mg/dL.
- An inadequate response to high-dose intravenous glucocorticoid treatment administered for at least 3 days as per local standard of care (including but not limited to pulses of 30 mg/kg prednisone on 3 consecutive days).
- High-dose intravenous glucocorticoids should not be lower than 2 mg/kg/day of prednisone equivalent in two divided doses (or at least 60 mg/day in patients of 30 kg or more). In case of rapid worsening of the patient’s condition and/or laboratory parameters, inclusion may occur within <3 days from starting high-dose intravenous glucocorticoids.
- Tocilizumab, tumour necrosis factor inhibitors, and canakinumab, if administered, had to be discontinued before emapalumab initiation.
- Informed consent provided by the patient (as required by local law) or by the patient’s legally authorised representative(s) with the assent of patients who are legally capable of providing it as applicable.
- Having received guidance on contraception for both male and female patients sexually active and having reached puberty:
  - Females of child-bearing potential required the use of highly effective contraceptive measures (failure rate of <1% per year)^a^ from screening until 6 months after receiving last dose of the investigational medicinal product.
  - Males with partners of child-bearing potential had to agree to take appropriate precautions (such as sexual abstinence, barrier contraception, or vasectomy) to avoid fathering a child from screening until 6 months after receiving last dose of the investigational medicinal product.

^a^ Highly effective contraceptive measures included: sexual abstinence; hormonal contraceptives: combination or progesterone only; intrauterine methods (intrauterine devices or systems); bilateral tubal occlusion; vasectomised partner.

**Exclusion criteria**

- Diagnosis of suspected or confirmed primary haemophagocytic lymphohistiocytosis or haemophagocytic lymphohistiocytosis consequent to a neoplastic disease.
- Active mycobacteria (typical and atypical), *Histoplasma capsulatum*, *Shigella*, *Salmonella*, *Campylobacter*, and *Leishmania* infections.
- Clinical suspicion of latent tuberculosis.
- Positive serology for human immunodeficiency virus antibodies.
- Presence of malignancy.
- Patients who had another concomitant disease or malformation severely affecting

cardiovascular, pulmonary, central nervous system, liver, or renal function that in the opinion of the investigator might have significantly affected likelihood to respond to treatment and/or assessment of emapalumab safety.

- History of hypersensitivity or allergy to any component of the study regimen.
- Receipt of bacillus Calmette-Guérin vaccine within 12 weeks prior to screening.
- Receipt of live or attenuated live vaccines within 6 weeks prior to screening.
- Pregnant or lactating female patients.

Abbreviations:

AOSD, adult-onset Still’s disease; MAS, macrophage activation syndrome; sJIA, systemic juvenile idiopathic arthritis

**Table S2. Signs and symptoms to be evaluated by the investigator while assessing the activity of MAS**

- Fever (greater than 38.0°C [100.4°F]).
- Skin rash.
- Haemorrhagic manifestations:
  - Skin bleeding (petecchiae, ecchymosis, or purpura) (the investigator had to choose among “stable”, “worsened”, “improved”, or “resolved”).
  - Mucosal bleeding (gut, respiratory) (the investigator had to choose among “present” or “absent”).
- Evidence of CNS involvement:
  - Clinical (headache, irritability, seizures, confusion, lethargy, coma) (the investigator had to choose among “stable”, “worsened”, “improved”, or “resolved”).
  - Cerebrospinal fluid abnormalities (cell count) (if lumbar puncture performed based on clinical indication).
- Respiratory function:
  - Oxygen support (the investigator had to choose among “present” or “absent”; if present, the investigator had to indicate how many liters of oxygen were required to maintain oxygen saturation).
  - Mechanical ventilation (the investigator had to choose among “present” or “absent”).
- Cardiac:
  - Pericarditis (if echocardiography was performed, results were to be reported).
  - Inotropic support.
- Kidney:
  - Ultrafiltration/dialysis.

Abbreviations:

CNS, central nervous system; MAS, macrophage activation syndrome

**Table S3. Total IFNγ, CXCL9, and sIL-2R levels during the study and the long-term follow-up study**

|  | **NI-0501-06 study** | | | | | | **Long-term follow-up (NI-0501-05 study)** | | |
| --- | --- | --- | --- | --- | --- | --- | --- | --- | --- |
|  | Day 0  (n=14) | Day 3  (n=14) | Day 7 (n=14) | Day 14  (n=14) | Day 28  (n=14) | Day 56  (n=14) | Day 100  (n=14) | 6 months  (n=10)^a^ | 12 months  (n=9)^a^ |
| **Total IFNγ** | | | | | | | | | |
| Median  (range) | <50  (<50–4510) | 2452  (66–37295) | 2456  (73–9886) | 1509  (59–20,744) | 1200  (<50–46,186) | 905  (<50–10,308) | 1207  (105–24,290) | 2177  (<50–12,675) | 594  (<50–4010) |
| Normal, n (%)^b^ | NA^c^ | 6 (43) | 7 (50) | 9 (64) | 10 (71) | 9 (64) | 10 (71) | 6 (60) | 7 (78) |
| **CXCL9** | | | | | | | | | |
| Median  (range) | 10884  (1020–98,121) | 1807  (598–21,963) | 555  (192–3044) | 217  (<80–1946) | 136  (<80–750) | 114  (<80–2296) | 142  (<80–1043) | 156  (<80–370) | 211  (111–2323) |
| Normal, n (%)^b^ | 0 (0) | 0 (0) | 1 (7) | 7 (50) | 12 (86) | 12 (86) | 12 (86) | 8 (80) | 6 (67) |
| **sIL-2R** | | | | | | | | | |
| Median  (range) | 5077  (1664–20,954) | 4669  (2646–36,481) | 3319  (1579–17,059) | 1756  (908–8090) | 1105  (571–3945) | 1098  (306–3432) | 840  (344–3122) | 853  (535–1803) | 904  (501–4019) |
| Normal, n (%)^b^ | 0 (0) | 0 (0) | 0 (0) | 1 (7) | 7 (50) | 7 (50) | 9 (64) | 6 (60) | 5 (56) |

^a^ In the long-term follow-up, the protocol required pharmacokinetic/pharmacodynamic sampling until emapalumab was undetectable in blood. Two patients with undetectable levels at Day 100 were therefore not sampled at 6 and 12 months, and 1 patient with undetectable levels at 6 months was not sampled at 12 months. One patient was not sampled at 6 months. One patient missed the 6- and 12- month visits of the long-term follow-up because of Covid-19-related travel restriction.

^b^ Below or equal to the 95^th^ percentile of healthy controls: 2084 pg/mL for total IFNγ, 271 pg/mL for CXCL9^1^ and 1071 pg/mL for sIL-2R.

^c^ At baseline, in the absence of emapalumab, total IFNγ consists only of free IFNγ. Therefore, the above indicated 95^th^ percentile for total IFNγ applies only to values obtained in the presence of emapalumab.

Abbreviations:

IFNγ, interferon-γ; sIL-2R, soluble interleukin-2 receptor

Reference: 1. Jacqmin P, Laveille C, Snoeck E, et al. Emapalumab in primary haemophagocytic lymphohistiocytosis and the pathogenic role of interferon gamma: A pharmacometric model-based approach. Br J Clin Pharmacol 2022;88(5):2128–2139.

**Table S4. Assessment of MAS remission at the end of the long-term follow-up study (12 months)**

Patients meeting MAS remission criteria: n=10

Patients not meeting MAS remission criteria: n=4

- One patient with MAS activity = 1.5
- One patient who stopped emapalumab after three administrations upon investigator’s assessment of remission, continued to have mild abnormalities only in LDH levels
- One patient had missing values for aspartate aminotransferase and LDH, with all other parameters of the criteria for MAS remission being met
- One patient missed the 6- and 12-month visits of the long-term follow-up because of Covid-19–related travel restrictions

Abbreviations: LDH, lactate dehydrogenase; MAS, macrophage activation syndrome

**Table S5. Inflammatory flares of underlying sJIA/AOSD and their treatment during the study and the long-term follow-up study**

| **NI-0501-06 study** | | |
| --- | --- | --- |
| **Patient ID** | **Flare and background treatment** | **Treatment of underlying sJIA/AOSD** |
| Patient 3 | - Anakinra (2 mg/kg/day) discontinued on study day 0 - CRP increase on study day 3 while on 2.5 mg/kg/day prednisone equivalent and MAS improving (first MAS remission achieved on study day 25) | - Anakinra re-introduced at 2 mg/kg/day on study day 4 |
| Patient 5 | - Anakinra (13 mg/kg/day) discontinued 11 days prior to start of emapalumab - CRP increase on study day 14 while on 2.5 mg/kg/day prednisone equivalent and MAS improving (first MAS achieved on study day 45) | - Anakinra re-introduced at 4 mg/kg/day on study day 14 |
| Patient 10 | - Anakinra (9 mg/kg) discontinued on study day -1 - CRP increase on study day 2 while on 2.1 mg/kg/day prednisone equivalent and MAS improvement (first MAS remission achieved on study day 23) | - Glucocorticoid pulses at 35 mg/kg/day prednisone equivalent on study day 4, 5, and 6 |
| Patient 13 | - CRP increase on study day 21 while receiving 1.5 mg/kg/day prednisone equivalent (emapalumab last dose on study day 24) and in MAS remission (first achieved on study day 21) | - Anakinra introduced at 2 mg/kg/day on study day 25, increased to 4 mg/kg/day on study day 26 |
| Patient 11 | - Persistently increased CRP from study day 0 while receiving 25 mg/kg/day prednisone equivalent and MAS improvement (first MAS remission achieved on study day 15) | - 30 mg/kg/day prednisone equivalent on study days 4 and 5. - Anakinra introduced on study day 6 at 2 mg/kg/day |
| Patient 8 | - CRP increase on study day 12 while on 0.4 mg/kg/day prednisone equivalent (last dose of emapalumab on study day 6) and MAS in resolution (only LDH > 1.5× ULN) | - Baricitinib introduced on study day 15 |

| **Long-term follow-up (NI-0501-05)** | | |
| --- | --- | --- |
| **Patient ID** | **Flare and background treatment** | **Treatment of underlying sJIA/AOSD** |
| Patient 1 | - Inflammatory flare of sJIA on study day 315 while on prednisone at 0.07 mg/kg/day and no MAS flare | - Glucocorticoid dose increased: methylprednisolone 0.9 mg/kg/day - Canakinumab introduced at 300 mg every 4 weeks |
| Patient 11 | - Inflammatory flare of sJIA on study day 233 while on anakinra at 1.8 mg/kg/day and off glucocorticoids and no MAS flare | - Methyl prednisolone pulses (1 g/day) for 3 consecutive days, switched to oral prednisone (0.9 mg/kg/day) and then tapered - Anakinra continued at 1.8 mg/kg/day |
| Patient 12 | - Inflammatory flare of sJIA on study day 189 while on anakinra at 14 mg/kg/day and off glucocorticoids | - Methylprednisolone pulses (1 g/day) for 2 days followed by prednisolone at 2.2 mg/kg/day - Subsequently tocilizumab (162 mg every 2 weeks) |
| Patient 14 | - Inflammatory flare of sJIA with interstitial lung disease on study day 320 while on prednisolone at 1.5 mg/kg/day, anakinra at 4.2 mg/kg/day and methotrexate 7.5 mg/week | - Tofacitinib introduced at 2.5 mg/day |

First MAS remission was the time of first fulfilment of MAS remission criteria

Flares of the underlying sJIA were identified based on adverse event reporting and on the rationale for change in medications or medication doses.

Abbreviations:

AOSD, adult-onset Still’s disease; CRP, C-reactive protein; LDH, lactate dehydrogenase; MAS, macrophage activation syndrome; sJIA, systemic juvenile idiopathic arthritis; ULN, upper limit of normal

**Table S6. TEAEs reported in ≥2 patients during the NI-0501-06 trial and the long-term follow-up study**

| **Event category** | **NI-0501-06 study**  **(up to week 8)**  (N=14) | **Long-term follow-up**  **(up to week 52)^a^**  (N=14) | |
| --- | --- | --- | --- |
| **TEAEs reported in ≥2 patients** | | | |
|  |  | Emapalumab levels at or above LLOQ | Emapalumab levels below LLOQ |
| No. of events | 23 | 9 | 5 |
| Rash | 4 | – | – |
| CMV infection reactivation | 3 | – | – |
| Diarrhoea | 4 | – | – |
| Hyperglycaemia | 3 | – | – |
| Hypertension | 3 | – | – |
| Headache | 4 | 1 | 1 |
| Injection site reaction | 2 | – | – |
| Viral URTI | – | 4 | 1 |
| sJIA flare | – | 3 | – |
| Cholelithiasis | – | – | 2 |
| Dental caries | – | 1 | 1 |

^a^ The protocol mandated that only serious AEs were to be reported after the level of emapalumab was undetectable. However, five non-serious AEs were also reported during this time and are included in the table.

Abbreviations:

AE adverse event; CMV, cytomegalovirus; LLOQ, lower limit of quantification; sJIA, systemic juvenile idiopathic arthritis; TEAE, treatment-emergent adverse event; URTI, upper respiratory tract infection
